# Preliminary Safety Analysis of Two Pilot Clinical Trials Involving Injections of Niagen®, Nicotinamide Riboside Chloride

**DOI:** 10.64898/2026.04.28.26352007

**Authors:** Yasmeen Nkrumah-Elie, Jun Kwon, Sierra Simpson, Rebecca Idoine, Jolie Mavoyan, Anne Russ, Jessie Cavanaugh, Liz Hawkins, Kayla Fuller, Jennifer Jaeger, Andrew Shao

**Affiliations:** ChromaDex Inc., a Niagen Bioscience, Inc. company, Los Angeles, California, USA; Verséa Health, Inc., Tampa, Florida, USA; Impact Health Medical FL PA, Orlando, Florida, USA; Nutraceuticals Research Institute, Huntsville, Alabama, USA

**Keywords:** nicotinamide riboside (NR), Niagen, nicotinamide adenine dinucleotide (NAD+), injection, intravenous, intramuscular, subcutaneous, clinical

## Abstract

Nicotinamide riboside (NR), an endogenous precursor to the essential coenzyme nicotinamide adenine dinucleotide (NAD+), is characterized as safe and effective at longitudinally elevating NAD+ in blood and tissues, when administered orally. Preclinical research on NR as an augmenter of NAD+ has demonstrated great promise in support of healthy aging, metabolic health, and several diseases, though clinical translation of these findings has been limited. Interest in alternative routes of administration of NR has increased in recent years and led to the development of pharmaceutical-grade NR for intravenous and injectable administration. Two separate Phase 1 pilot clinical trials were conducted evaluating NR via bolus injections. While the designs of the two studies are different, the similarities warrant combined presentation to note the similarities, particularly with regards to safety-related outcomes. Trial 1 involved 45 participants that were randomized to a 3x3 design, accounting for three interventions, placebo, NR, and NAD+ and three routes of administration, intramuscular (IM), intravenous (IV), and subcutaneous (SC), resulting in a 9-arm study (placebo IM, n=5; placebo IV, n=5; placebo SC, n=5; NR IM, n=6; NR IV, n=5; NR SC, n=4; NAD+ IM, n=4; NAD+ IV, n=5; and NAD+ SC, n=6). Participants were administered NR once daily for three days, followed by a 7-day washout period. In Trial 2 (n=39), the 2x2 study design incorporated 4-arms for phase 1, where the participants were randomized to 50 or 100 mg of NR, administered either IM or SC in-clinic for 3 consecutive days, followed by a 7-day washout (50 mg IM, n=7; IM, 100 mg IM, n=11; 50 mg SC, n=11; and 100 mg SC, n=10). Phase 2 of Trial 2 involved participants self-administering either 50 or 100 mg of NR subcutaneously. Safety assessments for both trials included vitals, blood biomarkers, participant reported outcomes regarding the experience, and adverse event monitoring. Participant retention for both studies was 100%, and the injections did not result in any attributable unexpected adverse events or experiences. The experiences described by the participants regarding the actual injection varied. In Trial 2, pain more than two minutes after the injection and muscle soreness and tightness were reported by 45.9 and 43.2% of the participants, respectively, regardless of the route of administration or dose. Vitals remained generally consistent throughout both trials, and reductions in systolic blood pressure observed in both trials should be evaluated in larger, properly powered studies. Blood chemistry biomarkers for both trials did not elicit treatment-related patterns. Both trials presented within-group reductions in hsCRP in the NR SC arms, however, given baseline imbalance and small samples size, this finding should be considered hypothesis-generating, only. Overall, in both trials, the interventions, regardless of route of administration were associated with some discomfort but were well tolerated and did not produce any concerning safety signals. It is recommended that comprehensive metabolic and inflammatory panels should continue to be employed in future studies and clinical settings to assess whether consistent patterns emerge in larger populations.

## 1. Introduction

Nicotinamide adenine dinucleotide (NAD+) is an essential coenzyme with numerous pivotal functions in cellular metabolic processes ^1^. In addition to its well-established role as a redox cofactor in cellular bioenergetics, NAD+ also functions as a critical substrate for protein families including sirtuins and poly-ADP-ribose polymerase (PARPs), which contribute to cellular processes including DNA repair^2^, immune signaling^3^, and post-translational protein modification^4,5^. Given these central roles, age-related declines in NAD+ concentrations across cells and tissues are of particular significance and may be further exacerbated by metabolic and environmental stressors such as overnutrition, excessive alcohol consumption, environmental toxicant exposure, and various disease states ^6–13^. The implications of NAD+ depletion are broad, and correlated to neurodegeneration, skeletal muscle dysfunction, and chronic inflammation^9,14,15^. Mechanistically, NAD+ and related changes in tissue NAD+ flux have been associated with the molecular hallmarks of aging, including mitochondrial dysfunction, telomere attrition, loss of proteostasis, genomic instability, cellular senescence, stem cell exhaustion, and chronic inflammation ^9,16–21^. Therefore, raising intracellular NAD+ has attracted scientific attention as a potential therapeutic target to mitigate age-associated molecular and functional decline ^9,14^.

The provision of intravenous (IV) NAD+ as a therapeutic and health-modifying strategy was first documented by O’Hollaren in 1961^22^. Then referred to as diphosphopyridine nucleotide (DPN), O’Hollaren reported beneficial uses of DPN in individuals with substance abuse disorders^22^, with subsequent investigations by an independent group of researchers exploring its application in schizophrenia ^23^. Since then, controlled clinical trials of parenteral NAD+ have remained sparse^24,25^. In contrast, IV NAD+’s use in wellness clinics for a variety of conditions has proliferated, largely based on anecdotal reports and clinical experience. Moreover, despite its long history of clinical use, the utility of exogenously administered NAD+, whether oral or parenterally, is limited by its phosphate groups, which preclude direct cellular uptake^26–28^. Instead, NAD+ undergoes extracellular degradation into its constituent components, such as nicotinamide (NAM) and nicotinamide riboside (NR), which subsequently enter cells through transporters present on the cellular membrane^29,30^. Interestingly, evidence suggests acute, temporal increases in extracellular NAD+ initiate an immune response or danger signal, resulting in a proliferation of proinflammatory molecules and toxicity conferred on T-cells ^31,32^. Such a response may explain the side effects associated with IV NAD+^33^.

Among strategies under investigation for NAD+ repletion, the pyridine nucleoside and NAD+ precursor, NR, has emerged as a promising candidate. A naturally occurring vitamin B3 derivative^34^, NR undergoes direct cellular entry and feeds into the salvage NAD+ biosynthetic pathway^35^, where nicotinamide riboside kinase (NRK) catalyzes its conversion to NAD+ - one of the three primary routes of NAD+ synthesis in mammals^36,37^. Comparative intravenous studies have demonstrated that exogenous NR is better tolerated^38,39^ and more effective at amplifying whole blood NAD+ than NAD+ itself ^39^. While clinical advancements in the IV administration of compounds purported to augment cellular NAD+ have continued, the use of injectable NR or NAD+ via subcutaneous (SC) or intramuscular (IM) routes remain largely unexplored, with no clinical studies in the published literature, to date, having systematically assessed these delivery approaches. If clinically translatable, the acute benefits of NR injections observed in preclinical models could have profound implications for the benefits of NAD+ on health outcomes^40–42^.

Oral NR supplementation has a reasonably well-characterized safety and efficacy profile, demonstrating the ability to increase whole blood NAD+ and augment NAD+ flux in tissues, in both clinical and preclinical models ^43–45^. In healthier populations, oral NR has yielded a number of modest but promising outcomes, such as improvements in mitochondrial biogenesis and muscle myoblast differentiation^46^, satellite cell proliferation^46^, metabolism changes^47^, reductions in inflammation ^45,48,49^ and substrate utilization efficiency^47^, though several of the robust metabolic and aging benefits observed preclinically have yet to be convincingly translated to human trials^50^. For example, while mitochondrial biogenesis was demonstrated in Lapatto et al. following five months of NR, other trials that have evaluated other indicators of mitochondrial biogenesis, such as PGC-1α protein, mtDNA copy number, citrate synthase activity, and OXPHOS complex content in human trials have had mixed results. Dollerup et al. (2018, 2020) and Remie et al. (2020) found no improvement in mitochondrial respiration or content in human muscle after oral NR^49,51,52^. These gaps and clinical inconsistencies, alongside the complexity of the bioavailability of oral NR – driven by its partial gastrointestinal and first-pass hepatic conversion to NA and NAM ^53,54^ – have prompted interest in other delivery strategies ^55,56^. Liu et al. 2018 demonstrated that intravenous administration of NR results in greater systemic availability compared to oral delivery, leading to direct increases in NAD+ levels in the liver, kidney, and muscle, with secondary elevations observed in the brain^57^. Parenteral administration bypasses gastrointestinal metabolism, which may improve plasma and tissue bioavailability of NR to enable more immediate and localized NAD+ augmentation and produce acute increases in NAD+ not observed with oral administration. Additionally, non-oral routes of NR administration may lead to more reliable, and robust increases in NAD+ at the tissue level^57^.

A key limitation in the clinical development of parenteral NR and NAD+ is the absence of repeat-dose toxicology studies to identify target organs of toxicity and establish the no observable adverse effects level (NOAEL). A recent median lethal dose (LD50) study of parenteral NR identified route-specific effects of NR administered via IV, IM, and SC administration, with estimated LD50s of >2000 mg/kg for SC and IM routes, and 1200-2000 mg/kg for the IV route ^58^. However, a notable limitation is that the LD50 estimates only provide a reference point for acute single-dose safety, and do not substitute for repeat-dose safety studies. Accordingly, the present pilot clinical studies were conservatively designed and exploratory in nature, given in the absence of foundational sub-chronic safety data and characterization.

To date, two pilot clinical trials have been conducted to evaluate the safety of Niagen® Plus (pharmaceutical grade nicotinamide riboside chloride) as an injectable product. For both studies, participants were administered the test product on days 1, 2, and 3, followed by a seven-day wash-out period and further observations on Day 10. The second study incorporated a 90-day at-home administration phase following the day 10 visit, with additional clinical visits conducted on days 40 and 100 to capture additional safety and exploratory efficacy endpoints. This paper only addresses the safety assessments from the two trials.

The first trial (Trial 1), “Absorption and Tolerability of Injectable Administration of Niagen®+, as Compared to NAD+,” was a nine-arm study that evaluated NAD+, NR, or placebo when administered as a bolus dose IM, IV, or SC, as shown in Table 1. The study was sponsored by Nutraceuticals Research Institute and funded by ChromaDex. The study was double-blind (administering clinician and participant), randomized trial.

**Table 1.** Trial 1 Study Design.

| Study Day | 1 | 2 | 3 |  | 10 |
| --- | --- | --- | --- | --- | --- |
| Clinical Site Visit # | 1 | 2 | 3 |  | 4 |
|  | Clinician-Administered Injections |  |  | 7-days | Post-Washout Follow-up |
| Placebo, IM | X | X | X | Washout | X |
| Placebo, IV | X | X | X |  | X |
| Placebo, SC | X | X | X |  | X |
| 100 mg NR, IM | X | X | X |  | X |
| 100 mg NR, IV | X | X | X |  | X |
| 100 mg NR, SC | X | X | X |  | X |
| 100 mg NAD<sup>+</sup>, IM | X | X | X |  | X |
| 100 mg NAD<sup>+</sup>, IV | X | X | X |  | X |
| 100 mg NAD<sup>+</sup>, SC | X | X | X |  | X |

The second trial (Trial 2), “Randomized, Open-label, Safety Study of Subcutaneous and Intramuscular Injections of Niagen® Plus,” sponsored by ChromaDex, Inc. was a 4-arm-washout-2-arm parallel design trial (Figure 1).

**Figure 1.**
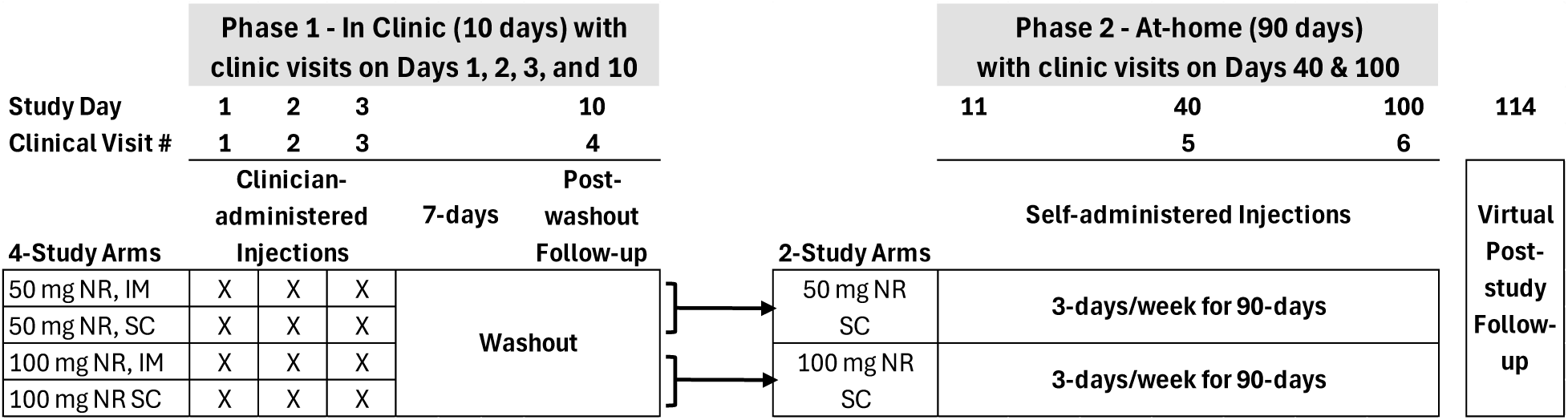
Trial 2 Study Design. Phase 1 of the study included 4-study arms, stratified by dose and route of administration for three consecutive-day administrations, followed by a 7-day washout. For phase 2 of the study, the participant groups were collapsed into two groups, based upon dose for at-home subcutaneous administration, 3 days per week for 90 days, with clinic visits on days 40 (Visit 5) and 100 (Visit 6). A virtual follow-up was conducted two weeks after the participants’ last injection.

## 2. Methods

### 2.1 Trial 1 Methods

#### 2.1.1. Trial 1 Ethics and Regulatory Authorization

The clinical trial, “Absorption and Tolerability of Injectable Administration of Niagen®+, as Compared to NAD+ (Trial 1),” was conducted in accordance with the Declaration of Helsinki, following Good Clinical Practice (GCP) guidelines. The clinical trial was administered by Nutraceuticals Research Institute (NRI) at their clinic in Huntsville, Alabama, USA. The study received authorization from the NRI Institutional Review Board, #1 (NIH OHRP registration number IRB00014538) to conduct the study under protocol number 24-09-1300 and was appropriately registered on ClinicalTrials.gov (NCT06919328). All participants provided written informed consent prior to study participation. Recruitment for the study began in October 2024 with last participant out in December 2024. Only de-identified data was provided to the authors, outside of the clinical team, to support data analysis.

#### 2.1.2. Trial 1 Design

Trial 1 was organized as a single-site, double-blind, randomized, placebo-controlled, prospective clinical pilot 3x3 trial with nine study arms. Participants were randomized to one of three interventions, placebo (vehicle only), NR, or NAD+. Participants were further randomized to a route of injectable administration, IM, IV, or SC, creating the nine treatment groups. The purpose of this study was to identify the continuity and variations between the three administration routes for NR and NAD+ and to serve as a pilot study to collect data for future powered studies. Accordingly, statistical significance was not expected in comparisons between the active treatments (NR and NAD+) and the placebo.

#### 2.1.3. Trial 1 Participants

Individuals of any gender, ages 40-65 with a body mass index of 25-34.9 kg/m^2^, with mild self-reported fatigue were deemed eligible for screening. Participants were recruited through an email database of over 15,000 individuals interested in participating in clinical trials. Eligible screened participants were required to meet the following additional inclusion criteria: 1) signed and dated informed consent form; 2) Stated willingness and demonstrated ability to comply with all study procedures, as well as availability for the duration of the study; 3) Live within 100 miles of the study site; 4) Sedentary behavior (defined as <20 minutes per day or <100 minutes per week of moderate to vigorous exercise); 5) Demonstrated above average fatigue as determined by the Fatigue Assessment Scale (FAS); 6) Females of reproductive potential agreed to use highly effective contraception for at least one month prior to screening, during study participation and one month after study completion and males agreed to use highly effective contraceptives during and 30-days after completion of the study; 7) Agreement to adhere to lifestyle consideration throughout the study, including a) a hydration standardization protocol 24 hours prior to each injection, b) refrain from dietary, lifestyle or exercise changes, c) immediate notification to the Primary Investigator (PI) if prescribed any new medications during the 10-day study period.

The following exclusion criteria were also employed: 1) Current diagnosis of a seizure disorder, diabetes or insulin resistance, kidney, liver, or cardiovascular disease, anemia, cancer, or Parkinson’s disease; 2) Any diagnosed chronic illness (pre-disease state acceptable); 3) Out of range phosphate levels at baseline; 4) BMI less than 25 or greater than or equal to 35; 5) Pregnancy, trying to conceive, or breastfeeding; 6) Known allergic reactions to any components of the intervention or related compounds, including any form of vitamin B3; 7) Positive COVID-19 test within 30 days of the study period; 8) Recent dramatic weight changes (10% change in body weight in the last 6 months); 9) Existing usage of a NAD+ or NAD+ precursor supplement in any form (oral, nasal, patch, or injection/IV administration), including a B-complex supplement, within 60 days of the Study; multivitamins were excluded; 10) Introducing a new medication, supplement, investigational drug, or other intervention (including lifestyle changes) within 60 days of the start of the study.

Participants were incentivized and compensated financially ($350) for completing all study requirements.

#### 2.1.4. Trial 1 Randomization and Blinding

Participants were randomized to one of the nine groups by adaptive randomization. For days 1, 2, and 3 of the intervention periods, participants were assigned to groups in the order they were listed to create a baseline for adaptive randomization. After this time, participant parameters and group assignments were entered into adaptive randomization software. The individual conducting the randomization, maintained the coding and was unblinded. The study participants and clinical staff remained blinded for the duration of the in-clinic portion of the study.

#### 2.1.5. Trial 1 Test Materials

NR was sourced as Niagen®Plus, pharmaceutical grade nicotinamide riboside chloride, obtained from W.R. Grace (South Haven, MI) for compounding and sterilization. NR and NAD+ were compounded at DCA Pharmacy (Franklin, TN) as single-use vials and maintained under refrigerated conditions until use. At time of administration, 100 mg of NR or NAD+ were dissolved in 2 ml of bacteriostatic water. The placebo control for the study was 2 ml of bacteriostatic water.

#### 2.1.6. Trial 1 Intervention

Consented participants were instructed to consume 60-120 oz of water the day prior to each clinic visit, and to fast for at least 8 hours, consuming only water, black coffee, or black tea during the fasting window. Each participant was clinically administered a bolus injection of either NR, NAD+, or placebo for three consecutive days, either IM, IV, or SC. After the third injection, a 7-day washout period commenced, and the participants returned on day 10 (clinic visit 4) for safety and efficacy assessments (Table 1).

#### 2.1.7. Trial 1 Safety Outcomes

The safety of the interventions was evaluated by monitoring vital signs (systolic blood pressure (SBP), diastolic blood pressure (DBP) heart rate (HR), respiratory rate (RR), oxygen saturation (SpO_2_), and temperature (Temp)), assessing blood biomarkers (glucose, insulin, high sensitivity C-reactive protein (hsCRP), serum viscosity, and erythrocyte sedimentation rate (ESR)), and documentation of adverse events reported by the participants and clinical staff.

#### 2.1.8. Trial 1 Outcomes to be reported elsewhere

The primary outcome of the study was to compare the subjective pain and discomfort experienced by participants from the various interventions using the Visual Analogue Scale (VAS) for pain, McGill Pain Questionnaire, and subjective, open-ended questions. Secondary outcomes included NAD+ measurements from dried blood spots and patient reported outcomes of NRI Fatigue Assessment Scale and 24-hour sleep-recall assessments.

#### 2.1.9. Trial 1 Qualitative Data Analysis

To supplement the structured safety assessments, participants were asked five open-ended questions at each injection visit to capture their subjective experience during and immediately following the injection. Responses were written by participants and reported verbatim. Following study completion, a post-hoc exploratory qualitative analysis was conducted on these qualitative data. This analysis was not pre-specified in the statistical analysis plan. Free-text responses were reviewed and inductively coded into descriptive symptom categories by members of the manuscript preparation team. Categories were generated from the data and were not derived from a standardized symptom inventory or validated instrument. Responses describing similar experiences were grouped under a single category label; where responses described distinct experiences, separate categories were retained. The frequency of coded responses was tabulated by intervention group and route of administration and is reported in Tables 4 and 5. Because this analysis is exploratory and post-hoc, and because inter-rater reliability was not assessed, findings should be interpreted as descriptive only. No inferential statistical comparisons between groups are supported by this analysis. The qualitative response data are presented to provide context for the structured safety outcomes and to generate hypotheses for future investigation.

#### 2.1.10. Trial 1 Statistical Approach

The intent-to-treat (ITT) population was designated as any participant that received one or more doses of the test substances, including the placebo. Continuous demographics and categorical variables were reported as means with standard error of the mean (SEM) or as group percentages. Vital signs were reported as the mean with SEM, as well as effect sizes with 95% confidence interval (CI). No formal between-arm hypothesis tests were pre-specified.

### 2.2 Trial 2 Methods

#### 2.2.1. Trial 2 Ethics and Regulatory Authorization

The clinical trial, “Randomized, Open-label, Safety Study of Subcutaneous and Intramuscular Injections of Niagen® Plus, (Trial 2),” was conducted in accordance with the Declaration of Helsinki, following Good Clinical Practice (GCP) guidelines. The clinical trial was administered by Impact Health Medical FL PA (Orlando, FL) at their clinic in Aventura, Florida, USA. The study received authorization from the Institutional Review Board of the Institute of Regenerative and Cellular Medicine IRB (HHS/OHRP IRB00009500) to conduct the study under protocol number NB-2025-01 (IRB approval number: IRCM-2023-368). The trial was registered on ClinicalTrials.gov (NCT07251608). All participants provided written informed consent prior to study participation. Recruitment for the study began in September 2025 with last participant out in February 2026. Only de-identified data was provided to the authors, outside of the clinical team, to support data analysis.

#### 2.2.2. Trial 2 Design

Trial 2 was designed and conducted as a single-site, prospective, two-phase, open-label, randomized clinical trial, comparing doses and routes of administration of injectable Niagen®, clinician- and self-administered, respectively. The first phase of the study consists of four-arms, 1) 50 mg NR administered IM, 2) 50 mg NR administered SC, 3) 100 mg NR administered IM, and 4) 100 mg NR administered SC. A goal of randomizing the participants 1:1:1:1 was attempted. The second phase consists of two arms, 1) 50 mg NR SC and 2) 100 mg NR SC. The purpose of this study was to characterize differences and consistencies across NR doses and routes of administration, and to generate pilot data to inform the powering for future studies. Accordingly, statistical significance was not expected when comparing NR doses and routes of administration to baseline values.

#### 2.2.3. Trial 2 Participants

Generally healthy adults of any gender, aged 18+, with no more than one, well-controlled, chronic condition were recruited (n=67) through IRB-approved informational documents, and community outreach. Outreach included the dissemination of digital documents at local community centers, wellness facilities, and public boards. Inclusion criteria included, 1) The demonstration of baseline fatigue as determined by a minimum FAS score (threshold prespecified in SAP); 2) NAD+ or NAD+ precursor naïve (including IV, IM, SC, or other administration route); 3) Non-anemic; 4) Willingness to adhere to lifestyle considerations and study procedures; 5) Ability to read English, and provide written informed consent; 6) Willingness to self-administer the study material, via subcutaneous injection for 90 days, and complete finger prick blood collections.

The participants’ lifestyle considerations required maintenance of typical diet from 2 days prior to Visit 1 until the completion of the assessments during Visit 4; maintenance of typical physical activity from 2 days prior to Visit 1 until the completion of Visit 6; avoidance of other NAD+ and NAD+ precursor products, with the exception of a multivitamin; adherence to a daily hydration requirement of ≥2L of water on the day prior to clinic visits; abstain from alcohol 24 hours prior to clinic visits; arrival at all clinic visits in a fasted state (minimum 8 hours, water only); conduct at-home assessments in a fasted state; refrain from smoking or vaping on clinic days; avoid the initiation of new medications unless medically necessary.

Exclusionary criteria included the following: 1) One or more uncontrolled chronic illnesses including diabetes, cardiovascular, liver, or kidney disease, or any form of cancer. An uncontrolled chronic illness was defined as any change to medication or other treatment modalities within the past 90 days; 2) More than one chronic disease under active treatment; 3) Any chronic disease, as determined by the primary investigator, that increases risk or confounds safety assessments; 4) Any acute illness within 14 days prior to Visit 1 (Day 1); 5) A cancer diagnosis within the past 5 years; 6) Anemia (defined as hemoglobin <100 g/L or other relevant hematologic abnormalities); 7) Current pregnancy or lactation, or unwillingness to use effective contraception if of childbearing potential; 8) Use of any NAD+ supplement, NAD+ precursor, or vitamin B_3_ product administered orally, nasally, transdermally, or by injection within the past 60 Days. NAD+ precursors and related compounds included niacin (NA), NAM, NR, nicotinamide mononucleotide (NMN), NAD+, NADH, inositol hexanicotinate, and apigenin. The exception was the use of a daily oral multivitamin that contained vitamin B_3_ (niacin/nicotinamide); 9) Hypersensitivity or allergy to NR, NA, other forms of vitamin B_3_/NAD+ precursors, or bacteriostatic water; 10) Significant aversion to needles or finger pricks; 11) Participation in another clinical intervention study within 90 Days (or 5 half-lives of the intervention, whichever is longer) prior to Visit 1 (Day 1); 12) Any other condition rendering the participant unsuitable, as determined by the investigator; 13) Excessive alcohol use, defined as 4 or more drinks on a single occasion, or illicit drug use that would prevent adherence to the protocol as determined by the investigator.

Participants were not financially compensated for their participation in the study. They were provided with blood biomarker and other results not reported in this manuscript upon study completion. In cases of clinically concerning laboratory findings, participants were notified prior to study completion and provided with their lab results.

#### 2.2.4. Trial 2 Randomization and Blinding

Randomization was conducted using a computer-generated allocation schedule with concealed assignment until the time of allocation. Blinding was not incorporated into the study design or data analysis plan. Given the community-based recruitment approach, some participants were acquainted and may have discussed aspects of the study, including dosing, route of administration, and perceived outcomes, which represents a potential source of bias.

#### 2.2.5. Trial 2 Investigational Product

Niagen®Plus, pharmaceutical grade NR chloride, was prepared and compounded as a sterile powder for reconstitution containing 500 mg per multi-use vial by Wells Pharma (Houston, TX). The study material was stored at 2–8 °C and protect from light when not in use. NR was reconstituted by the clinician staff with bacteriostatic water for the individual participants. Vials were only used for up to 28 days, per FDA guidelines for multiuse vial products.

#### 2.2.6. Trial 2 Intervention

Consented participants were instructed to drink >2 liters of water the day before clinic visits and to arrive fasted. Participants were randomized to the four-arms during the first phase, 1) 50 mg IM, 2) 50 mg SC, 3) 100 mg IM, and 4) 100 mg SC. NR administration in phase 1 of the study was conducted by a qualified clinician for three consecutive days (Visits 1-3; Days 1-3). Participants were administered NR in the deltoid, upper outer quadrant for IM and in the abdomen, below the ribs and above the iliac crest, ≥5 cm from umbilicus for SC, rotating sides with each administration. Following the 7-day washout, during clinic visit 4 (Day 10), the 4 study arms from phase 1 were collapsed into 2 groups for phase 2, 50 mg SC or 100 mg SC for the remaining 90-days of the study (Figure 1). The dose for the participants did not change between the study phases, however, participants that were administered NR IM, were changed to self-administering the product SC. Participants were instructed on how to conduct SC self-administration in the abdomen or upper thigh, rotating sites. Participants agreed to inject themselves 3 days per week, and for their last injection prior to clinic visits 5 (Day 40) and 6 (Day 100) to administer the product 24 hours prior to their arrival.

#### 2.2.7. Trial 2 Safety Outcomes

To assess the primary outcome of the study, the safety of NR injections for 100 days, vital signs, blood biomarkers, and documentation of adverse events described by study participants and the clinical staff were collected. Vital signs (SBP, DBP, HR, RR, SpO_2_, and temp) were conducted using calibrated devices, in a standardized (seated) position. For the first NR administration, vitals and adverse were collected at baseline, 15-, 60-, 90-, and 180 minutes post-injection. For visits 2 and 3, they were collected before (visit baseline) and 90-minutes post injection, and for visits 5 and 6, they were collected at baseline only. Venous blood and plasma collections to support a comprehensive metabolic panel (CMP), complete blood count, homocysteine, and hsCRP were conducted at baseline for visits 1, 4, 5, and 6, and at 90-minutes post injection for visits 1-3. Additionally, subjective experiences regarding tolerability, pain, and soreness were collected.

#### 2.2.8. Trial 2 Outcomes to be reported elsewhere

The secondary outcome of the pharmacodynamics of a single injected dose of NR on capillary whole blood NAD+, as measured by dried blood spots, and exploratory outcomes of participant reported outcomes (PRO) for fatigue, sleep, quality of life, skin quality, and overall experience, as well mitochondrial efficiency and urinary oxidative stress markers, will be explored in future manuscripts.

#### 2.2.9. Statistical Approach - complete

The Intent-to-Treat and safety sets included all participants that received one or more injections. Statistical analysis for vitals and blood biomarkers incorporated two-group comparisons (e.g., V1 (Visit 1) Baseline vs V1 90 min): Paired t-test (scipy.stats.ttest_rel) on matched subjects. Time course comparisons (3+ timepoints, e.g., V1→V6 or V1/V2/V3 90 min) involved One-way ANOVA (scipy.stats.f_oneway) for omnibus test, followed by Tukey HSD post hoc (statsmodels.stats.multicomp.pairwise_tukeyhsd) for pairwise comparisons. Data were represented as averages ±SEM, unless otherwise noted.

## 3. Results

### 3.1. Trial 1 Results

#### 3.1.1 Trial 1 Participants

A total of 299 participants within 100 miles of Huntsville, Alabama were contacted regarding Trial 1, with 137 screened for qualification; 46 participants met the inclusion criteria and provided informed consent. Forty-five participants were randomized for intent-to-treat (ITT), and all participants completed the required three injections and 7-day follow-up, providing a per protocol study population of N=45. The study was a parallel 3X3 design for intervention and route of administration, thus resulting in 9-arms to the study (Figure 2).

**Figure 2.**
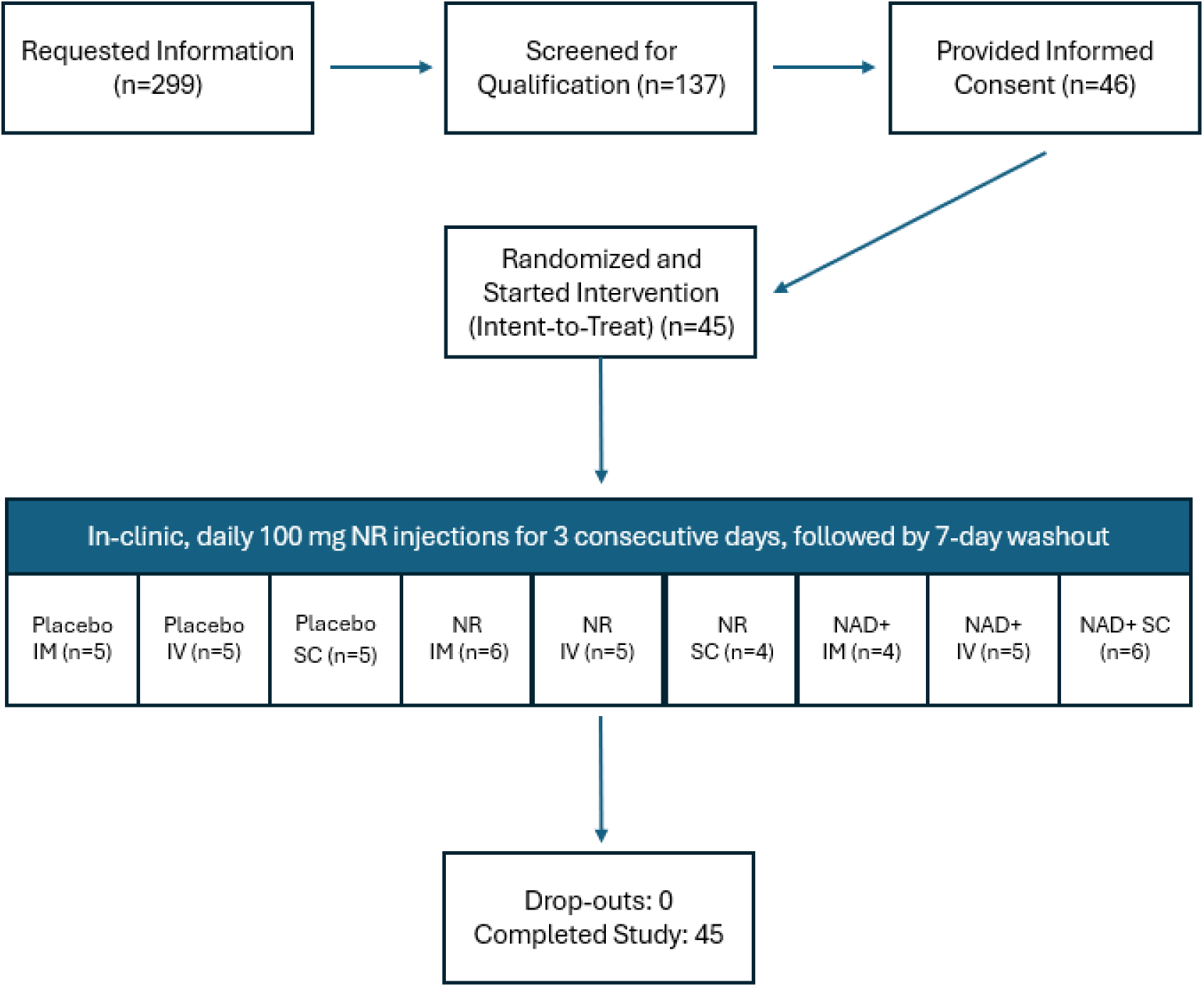
Trial 1 Participant Flow Diagram

#### 3.1.2 Trial 1 Demographics

The participants ranged in age from 40-64 (average 50.44 ± 1.02), skewed more female (71%) to male (29%), mostly identified as white (87%), presented with overweight or obesity (BMI: 29.4 kg/m2 ± 0.43), and had mild- to-moderate fatigue at screening (28.2 ± 0.83). The goal of 5 participants for each of the 9 groups was not met, though nearly achieved; six participants were in the NR IM and NAD+ SC groups, 4 participants were in the NR SC and NAD+ IM groups, and the remaining 5 groups each had 5 participants. There were no statistically significant differences across the nine groups for age, BMI, and baseline fatigue assessment scale (FAS) in Trial 1 (Table 2).

**Table 2.**
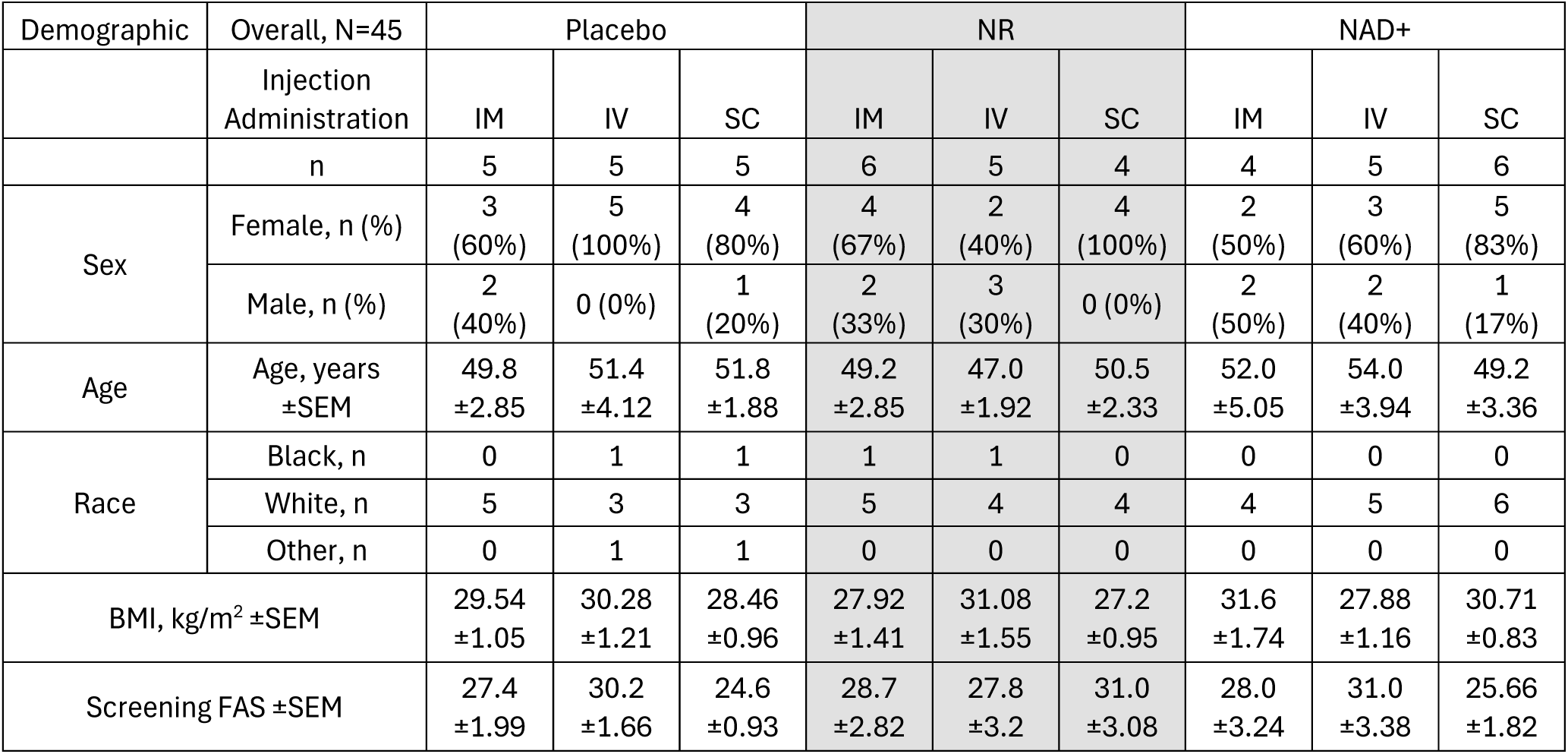
Trial 1 Baseline Demographics. Values for age, BMI, and baseline FAS are expressed as mean ± SEM.

#### 3.1.3. Trial 1 Vitals

Vitals were assessed before and after each injection to determine if there was an acute response to the intervention. Vitals remained in normal ranges for the duration of the study. SBP from the NR IM group was the only vital parameter with non-overlapping confidence intervals between baseline and post injection 3 (124.7-142 vs 113.5-122.5 respectively).

There were no significant changes in diastolic blood pressure, heart rate, respiratory rate, or temperature for any group over the course of Trial 1 (Table 3), suggesting that within this study population, the vitals remained stable and within normal ranges, consistent with the baseline data, indicating no safety concerns.

**Table 3.** Trial 1 Vitals. Vitals are presented as averages, plus or minus the standard error of the mean (SEM), as well as the 95% confidence interval (CI) for placebo, NR, and NAD groups, via route of administration.

| Vital Parameter | Treatment Group |  | Placebo |  |  |  |  | NR |  |  |  |  | NAD |  |  |  |  |
| --- | --- | --- | --- | --- | --- | --- | --- | --- | --- | --- | --- | --- | --- | --- | --- | --- | --- |
|  |  |  | Baseline | 90-min Post Inj 1 | 90 min Post Inj 2 | 90 min Post Inj 3 | After 7-day Washout | Baseline | 90-min Post Inj 1 | 90 min Post Inj 2 | 90 min Post Inj 3 | After 7-day Washout | Baseline | 90-min Post Inj 1 | 90 min Post Inj 2 | 90 min Post Inj 3 | After 7-day Washout |
| SBP, mmHG | IM | Avg | 126.6 | 127.2 | 129.2 | 126.2 | 124.4 | 133.3 | 129.2 | 130.8 | 118.0 | 124.0 | 132.0 | 128.5 | 132.3 | 125.5 | 127.5 |
|  |  | ±SEM | 3.458 | 1.96 | 2.888 | 3.499 | 3.723 | 3.363 | 5.741 | 4.362 | 1.732 | 5.768 | 2.483 | 3.096 | 3.351 | 1.848 | 7.03 |
|  |  | n | 5 | 5 | 5 | 5 | 5 | 6 | 6 | 6 | 6 | 6 | 4 | 4 | 4 | 4 | 4 |
|  |  | 95% CI | 117.0-136.2 | 121.8-132.6 | 121.2-137.2 | 116.5-135.9 | 114.1-134.7 | 124.7-142.0 | 114.4-143.9 | 119.6-142.0 | 113.5-122.5 | 109.2-138.8 | 124.1-139.9 | 118.6-138.4 | 121.6-142.9 | 119.6-131.4 | 105.1-149.9 |
|  | IV | Avg | 128.4 | 124.2 | 120 | 122.4 | 122 | 132.6 | 134.4 | 128.2 | 128.2 | 121 | 127 | 127.2 | 118.4 | 121.2 | 119.4 |
|  |  | ±SEM | 4.874 | 3.089 | 3.633 | 3.6 | 3.962 | 3.919 | 2.462 | 4.883 | 1.068 | 3.317 | 2 | 2.396 | 2.315 | 2.107 | 7.386 |
|  |  | n | 5 | 5 | 5 | 5 | 5 | 5 | 5 | 5 | 5 | 5 | 5 | 5 | 5 | 5 | 5 |
|  |  | 95% CI | 114.9-141.9 | 115.6-132.8 | 109.9-130.1 | 112.4-132.4 | 111.0-133.0 | 121.7-143.5 | 127.6-141.2 | 114.6-141.8 | 125.2-131.2 | 111.8-130.2 | 121.4-132.6 | 120.5-133.9 | 112.0-124.8 | 115.3-127.1 | 98.89-139.9 |
|  | SC | Avg | 130.2 | 123.8 | 127.8 | 126.6 | 128.2 | 139 | 139.5 | 127.5 | 133.5 | 126.8 | 132 | 136.2 | 133.5 | 126.3 | 121 |
|  |  | ±SEM | 8.015 | 3.184 | 6.666 | 3.156 | 7.144 | 4.021 | 4.941 | 4.031 | 7.24 | 3.146 | 4.626 | 4.453 | 4.161 | 5.69 | 1.528 |
|  |  | n | 5 | 5 | 5 | 5 | 5 | 4 | 4 | 4 | 4 | 4 | 6 | 6 | 6 | 6 | 6 |
|  |  | 95% CI | 107.9-152.5 | 115.0-132.6 | 109.3-146.3 | 117.8-135.4 | 108.4-148.0 | 126.2-151.8 | 123.8-155.2 | 114.7-140.3 | 110.5-156.5 | 116.7-136.8 | 120.1-143.9 | 124.7-147.6 | 122.8-144.2 | 111.7-141.0 | 117.1-124.9 |
| DBP, mmHG | IM | Avg | 80 | 84.8 | 81.6 | 79.6 | 79.6 | 86.83 | 84.83 | 83 | 77.5 | 79.83 | 81.75 | 79 | 84.75 | 80.75 | 84.25 |
|  |  | ±SEM | 2.983 | 1.356 | 4.118 | 4.155 | 0.5099 | 2.4 | 3.103 | 3.54 | 2.377 | 4.881 | 2.594 | 4.416 | 2.75 | 3.902 | 6.969 |
|  |  | n | 5 | 5 | 5 | 5 | 5 | 6 | 6 | 6 | 6 | 6 | 4 | 4 | 4 | 4 | 4 |
|  |  | 95% CI | 71.72-88.28 | 81.03-88.57 | 70.17-93.03 | 68.07-91.13 | 78.18-81.02 | 80.66-93.00 | 76.86-92.81 | 73.90-92.10 | 71.39-83.61 | 67.29-92.38 | 73.49-90.01 | 64.95-93.05 | 76.00-93.50 | 68.33-93.17 | 62.07-106.4 |
|  | IV | Avg | 76.8 | 79.4 | 77.2 | 77.4 | 78.6 | 86.8 | 81.8 | 84.8 | 82.4 | 84.6 | 85 | 83 | 80 | 78.8 | 82 |
|  |  | ±SEM | 0.5831 | 3.108 | 2.634 | 1.47 | 2.638 | 4.079 | 3.137 | 2.746 | 3.059 | 1.631 | 2.049 | 3.146 | 4.438 | 4.42 | 3.975 |
|  |  | n | 5 | 5 | 5 | 5 | 5 | 5 | 5 | 5 | 5 | 5 | 5 | 5 | 5 | 5 | 5 |
|  |  | 95% CI | 75.18-78.42 | 70.77-88.03 | 69.89-84.51 | 73.32-81.48 | 71.28-85.92 | 75.47-98.13 | 73.09-90.51 | 77.18-92.42 | 73.91-90.89 | 80.07-89.13 | 79.31-90.69 | 74.26-91.74 | 67.68-92.32 | 66.53-91.07 | 70.96-93.04 |
|  | SC | Avg | 83.20 | 81.80 | 82.00 | 74.20 | 86.80 | 83.00 | 83.50 | 80.25 | 80.00 | 83.00 | 85.17 | 83.00 | 91.33 | 84.00 | 78.00 |
|  |  | ±SEM | 3.967 | 1.53 | 4 | 2.223 | 4.116 | 1.732 | 3.227 | 7.284 | 5.701 | 1.581 | 5.455 | 5.768 | 5.414 | 3.624 | 1.633 |
|  |  | n | 5 | 5 | 5 | 5 | 5 | 4 | 4 | 4 | 4 | 4 | 6 | 6 | 6 | 6 | 6 |
|  |  | 95% CI | 72.18-94.22 | 77.55-86.05 | 70.89-93.11 | 68.03-80.37 | 75.37-98.23 | 77.49-88.51 | 73.23-93.77 | 57.07-103.4 | 61.86-98.14 | 77.97-88.03 | 71.14-99.19 | 68.17-97.83 | 77.42-105.3 | 74.68-93.32 | 73.80-82.20 |
| HR, bpm | IM | Avg | 80.6 | 76.4 | 76 | 77.4 | 84 | 90.5 | 80.67 | 77.67 | 80.17 | 83.17 | 80.75 | 79.75 | 74 | 72.25 | 86.25 |
|  |  | ±SEM | 1.939 | 2.205 | 3.082 | 4.25 | 3.912 | 4.349 | 3.095 | 1.667 | 2.845 | 5.282 | 3.772 | 3.75 | 3.764 | 5.935 | 4.661 |
|  |  | n | 5 | 5 | 5 | 5 | 5 | 6 | 6 | 6 | 6 | 6 | 4 | 4 | 4 | 4 | 4 |
|  |  | 95% CI | 75.22-85.98 | 70.28-82.52 | 67.44-84.56 | 65.60-89.20 | 73.14-94.86 | 79.32-101.7 | 72.71-88.62 | 73.38-81.95 | 72.85-87.48 | 69.59-96.74 | 68.75-92.75 | 67.82-91.68 | 62.02-85.98 | 53.36-91.14 | 71.42-101.1 |
|  | IV | Avg | 80.00 | 75.80 | 78.00 | 85.00 | 82.40 | 76.00 | 78.40 | 75.80 | 76.60 | 76.60 | 78.20 | 76.20 | 75.00 | 78.20 | 82.40 |
|  |  | ±SEM | 3.479 | 3.693 | 6.542 | 4.427 | 5.819 | 2.864 | 2.502 | 4.984 | 3.027 | 3.265 | 3.513 | 2.035 | 1.549 | 1.985 | 4.445 |
|  |  | n | 5 | 5 | 5 | 5 | 5 | 5 | 5 | 5 | 5 | 5 | 5 | 5 | 5 | 5 | 5 |
|  |  | 95% CI | 70.34-89.66 | 65.55-86.05 | 59.84-96.16 | 72.71-97.29 | 66.24-98.56 | 68.05-83.95 | 71.45-85.35 | 61.96-89.64 | 68.20-85.00 | 67.54-85.66 | 68.45-87.95 | 70.55-81.85 | 70.70-79.30 | 72.69-83.71 | 70.06-94.74 |
|  | SC | Avg | 71.6 | 76.4 | 79.6 | 87.2 | 76.6 | 76.5 | 73.75 | 73.5 | 74.75 | 76.25 | 76.83 | 80.67 | 80.83 | 76.67 | 81.83 |
|  |  | ±SEM | 4.032 | 4.874 | 3.982 | 1.655 | 3.124 | 5.867 | 7.983 | 5.605 | 1.887 | 6.588 | 3.953 | 4.455 | 7.323 | 3.612 | 4.061 |
|  |  | n | 5 | 5 | 5 | 5 | 5 | 4 | 4 | 4 | 4 | 4 | 6 | 6 | 6 | 6 | 6 |
|  |  | 95% CI | 60.40-82.80 | 62.87-89.93 | 68.54-90.66 | 82.60-91.80 | 67.93-85.27 | 57.83-95.17 | 48.34-99.16 | 55.66-91.34 | 68.74-80.76 | 55.29-97.21 | 66.67-87.00 | 69.22-92.12 | 62.01-99.66 | 67.38-85.95 | 71.39-92.27 |

Table 3 continued
| Vital<br>Parameter | Treatment<br>Group |  | Placebo |  |  |  |  | NR |  |  |  |  | NAD |  |  |  |  |
| --- | --- | --- | --- | --- | --- | --- | --- | --- | --- | --- | --- | --- | --- | --- | --- | --- | --- |
|  |  |  | Baseline | 90-min<br>Post Inj 1 | 90 min<br>Post Inj 2 | 90 min<br>Post Inj 3 | After 7-day<br>Washout | Baseline | 90-min<br>Post Inj 1 | 90 min<br>Post Inj 2 | 90 min<br>Post Inj 3 | After 7-day<br>Washout | Baseline | 90-min<br>Post Inj 1 | 90 min<br>Post Inj 2 | 90 min<br>Post Inj 3 | After 7-day<br>Washout |
| RR, brpm | IM | Avg | 16.4 | 16.8 | 16.8 | 16.2 | 16.6 | 16.33 | 16.67 | 16.33 | 16.5 | 17 | 16.25 | 16 | 16.5 | 16.25 | 16.25 |
|  |  | ±SEM | 0.4 | 0.4899 | 0.4899 | 0.2 | 0.4 | 0.3333 | 0.4216 | 0.3333 | 0.3416 | 0.4472 | 0.25 | 0 | 0.5 | 0.25 | 0.25 |
|  |  | n | 5 | 5 | 5 | 5 | 5 | 6 | 6 | 6 | 6 | 6 | 4 | 4 | 4 | 4 | 4 |
|  |  | 95% CI | 15.29-17.51 | 15.44-18.16 | 15.44-18.16 | 15.64-16.76 | 15.49-17.71 | 15.48-17.19 | 15.58-17.75 | 15.48-17.19 | 15.62-17.38 | 15.85-18.15 | 15.45-17.05 | 16.00-16.00 | 14.91-18.09 | 15.45-17.05 | 15.45-17.05 |
|  | IV | Avg | 16.8 | 17 | 16.2 | 16.4 | 17 | 16.8 | 17 | 16 | 16.6 | 16 | 17 | 16.4 | 16.4 | 17.2 | 16.6 |
|  |  | ±SEM | 0.3742 | 0.3162 | 0.2 | 0.2449 | 0.4472 | 0.4899 | 0.4472 | 0 | 0.4 | 0 | 0.4472 | 0.4 | 0.4 | 0.4899 | 0.4 |
|  |  | n | 5 | 5 | 5 | 5 | 5 | 5 | 5 | 5 | 5 | 5 | 5 | 5 | 5 | 5 | 5 |
|  |  | 95% CI | 15.76-17.84 | 16.12-17.88 | 15.64-16.76 | 15.72-17.08 | 15.76-18.24 | 15.44-18.16 | 15.76-18.24 | 16.00-16.00 | 15.49-17.71 | 16.00-16.00 | 15.76-18.24 | 15.29-17.51 | 15.29-17.51 | 15.84-18.56 | 15.49-17.71 |
|  | SC | Avg | 16.8 | 16.2 | 16 | 16.4 | 16.6 | 16.75 | 16.5 | 16.25 | 16 | 16.75 | 16.83 | 16.5 | 16.33 | 16.67 | 16.5 |
|  |  | ±SEM | 0.4899 | 0.2 | 0 | 0.4 | 0.4 | 0.4787 | 0.5 | 0.25 | 0 | 0.4787 | 0.654 | 0.3416 | 0.3333 | 0.4216 | 0.3416 |
|  |  | n | 5 | 5 | 5 | 5 | 5 | 4 | 4 | 4 | 4 | 4 | 6 | 6 | 6 | 6 | 6 |
|  |  | 95% CI | 15.44-18.16 | 15.64-16.76 | 16.00-16.00 | 15.29-17.51 | 15.49-17.71 | 15.23-18.27 | 14.91-18.09 | 15.45-17.05 | 16.00-16.00 | 15.23-18.27 | 15.15-18.51 | 15.62-17.38 | 15.48-17.19 | 15.58-17.75 | 15.62-17.38 |
| Temp, °F | IM | Avg | 98.36 | 98.14 | 98.2 | 97.68 | 98.04 | 98.02 | 97.93 | 98.05 | 98.17 | 98.12 | 98.4 | 97.75 | 98.33 | 97.95 | 97.68 |
|  |  | ±SEM | 0.1965 | 0.103 | 0.08367 | 0.1463 | 0.1208 | 0.147 | 0.1892 | 0.08466 | 0.1358 | 0.174 | 0.255 | 0.2901 | 0.1601 | 0.0866 | 0.1493 |
|  |  | n | 5 | 5 | 5 | 5 | 5 | 6 | 6 | 6 | 6 | 6 | 4 | 4 | 4 | 4 | 4 |
|  |  | 95% CI | 97.81-98.91 | 97.85-98.43 | 97.97-98.43 | 97.27-98.09 | 97.70-98.38 | 97.64-98.39 | 97.45-98.42 | 97.83-98.27 | 97.82-98.52 | 97.67-98.56 | 97.59-99.21 | 96.83-98.67 | 97.82-98.83 | 97.67-98.23 | 97.20-98.15 |
|  | IV | Avg | 98 | 97.94 | 97.84 | 98 | 98.06 | 98.18 | 97.88 | 98.34 | 98.06 | 97.98 | 97.9 | 98.16 | 98.46 | 97.96 | 98 |
|  |  | ±SEM | 0.05477 | 0.1122 | 0.413 | 0.1703 | 0.147 | 0.153 | 0.1715 | 0.2112 | 0.09274 | 0.18 | 0.2881 | 0.06 | 0.1806 | 0.1166 | 0.06325 |
|  |  | n | 5 | 5 | 5 | 5 | 5 | 5 | 5 | 5 | 5 | 5 | 5 | 5 | 5 | 5 | 5 |
|  |  | 95% CI | 97.85-98.15 | 97.63-98.25 | 96.69-98.99 | 97.53-98.47 | 97.65-98.47 | 97.76-98.60 | 97.40-98.36 | 97.75-98.93 | 97.80-98.32 | 97.48-98.48 | 97.10-98.70 | 97.99-98.33 | 97.96-98.96 | 97.64-98.28 | 97.82-98.18 |
|  | SC | Avg | 97.54 | 97.88 | 98.12 | 97.86 | 98.14 | 97.88 | 98.25 | 97.9 | 97.93 | 98.05 | 98.15 | 98.22 | 97.9 | 98.13 | 97.78 |
|  |  | ±SEM | 0.1939 | 0.2956 | 0.2634 | 0.1435 | 0.1208 | 0.3065 | 0.3175 | 0.2944 | 0.1887 | 0.1258 | 0.1384 | 0.1222 | 0.4107 | 0.08819 | 0.1851 |
|  |  | n | 5 | 5 | 5 | 5 | 5 | 4 | 4 | 4 | 4 | 4 | 6 | 6 | 6 | 6 | 6 |
|  |  | 95% CI | 97.00-98.08 | 97.06-98.70 | 97.39-98.85 | 97.46-98.26 | 97.80-98.48 | 96.90-98.85 | 97.24-99.26 | 96.96-98.84 | 97.32-98.53 | 97.65-98.45 | 97.79-98.51 | 97.90-98.53 | 96.84-98.96 | 97.91-98.36 | 97.31-98.26 |

**Table. 4.**
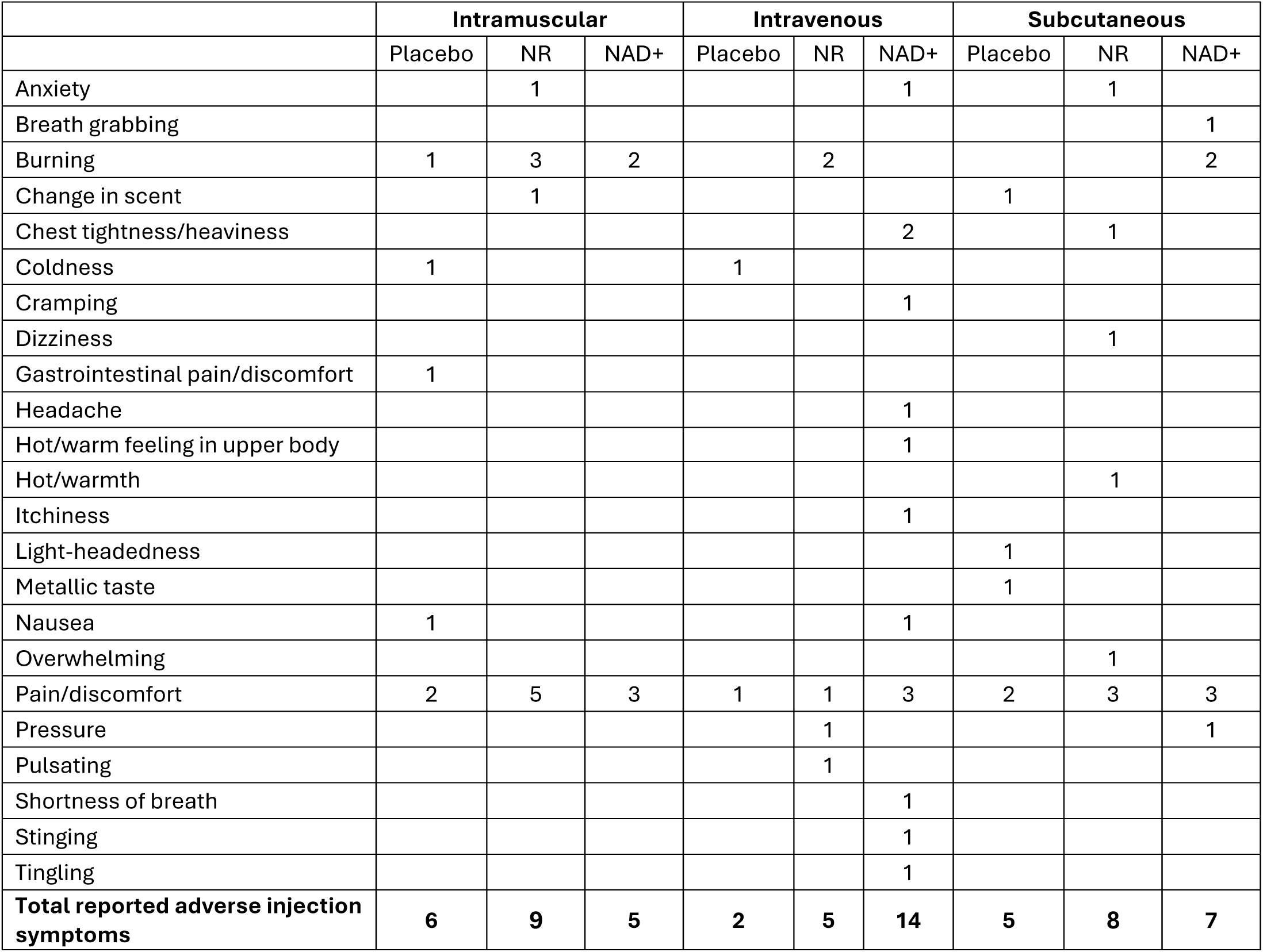
Summation of free-text symptom descriptions reported during injection, categorized post-hoc.

**Table 5.** Summation of free-text symptom descriptions reported 5-minutes post-injection, categorized post-hoc.

|  | Intramuscular |  |  | Intravenous |  |  | Subcutaneous |  |  |
| --- | --- | --- | --- | --- | --- | --- | --- | --- | --- |
|  | Placebo | NR | NAD+ | Placebo | NR | NAD+ | Placebo | NR | NAD+ |
| A strong radiate to fingers |  |  |  |  |  |  |  |  | 1 |
| Burning |  |  | 1 |  |  |  |  |  | 1 |
| Buzzing |  |  |  |  |  |  | 1 |  |  |
| Chest tightness/heaviness |  |  |  |  |  | 1 |  |  |  |
| Coldness |  |  |  |  |  |  | 1 |  |  |
| Fuzzy headed |  | 1 |  |  |  |  |  |  |  |
| Gastrointestinal pain/discomfort |  |  |  |  |  | 1 |  |  |  |
| Headache |  | 1 | 1 |  |  |  |  |  |  |
| Itchiness |  |  |  |  |  | 2 |  | 1 |  |
| Muscle tightness |  | 1 |  |  |  |  |  |  |  |
| Nausea |  |  | 1 |  |  | 1 |  |  |  |
| Neck tightness |  |  |  |  |  |  |  | 1 |  |
| Pain/discomfort | 2 | 3 | 3 | 1 | 1 | 1 | 2 | 2 | 3 |
| Paralysis/can't move |  |  |  |  |  | 1 |  |  | 1 |
| Pressure |  |  |  | 1 |  |  |  |  | 1 |
| Pulsating at injection site |  |  |  |  |  |  |  | 1 |  |
| Reductions in uncomfortable symptoms; return to normal |  |  |  |  |  | 1 |  |  |  |
| Return to normal | 1 |  |  |  |  |  |  | 1 |  |
| Shortness of breath |  |  |  |  |  | 1 |  |  |  |
| Tightness |  |  |  | 1 |  |  |  |  |  |
| Tingling |  |  |  |  | 1 |  |  |  |  |
| **Total reported adverse experiences post injection** | **2** | **6** | **6** | **3** | **2** | **8** | **4** | **5** | **7** |

#### 3.1.4. Trial 1 Blood Biomarkers (Glucose, insulin, inflammatory markers)

Blood biomarkers for fasted blood glucose, serum insulin, and clinical inflammatory markers were assessed throughout the trial. A baseline comprehensive metabolic panel and complete blood count (CBC) were assessed at baseline, only (data not shown), but were not assessed throughout the study to monitor acute changes in safety blood biomarkers. The causative relationship between the directional changes in fasted glucose and insulin are not apparent, and may be related to individual variability in these parameters (Figures 3 and 4).

**Figure 3.**
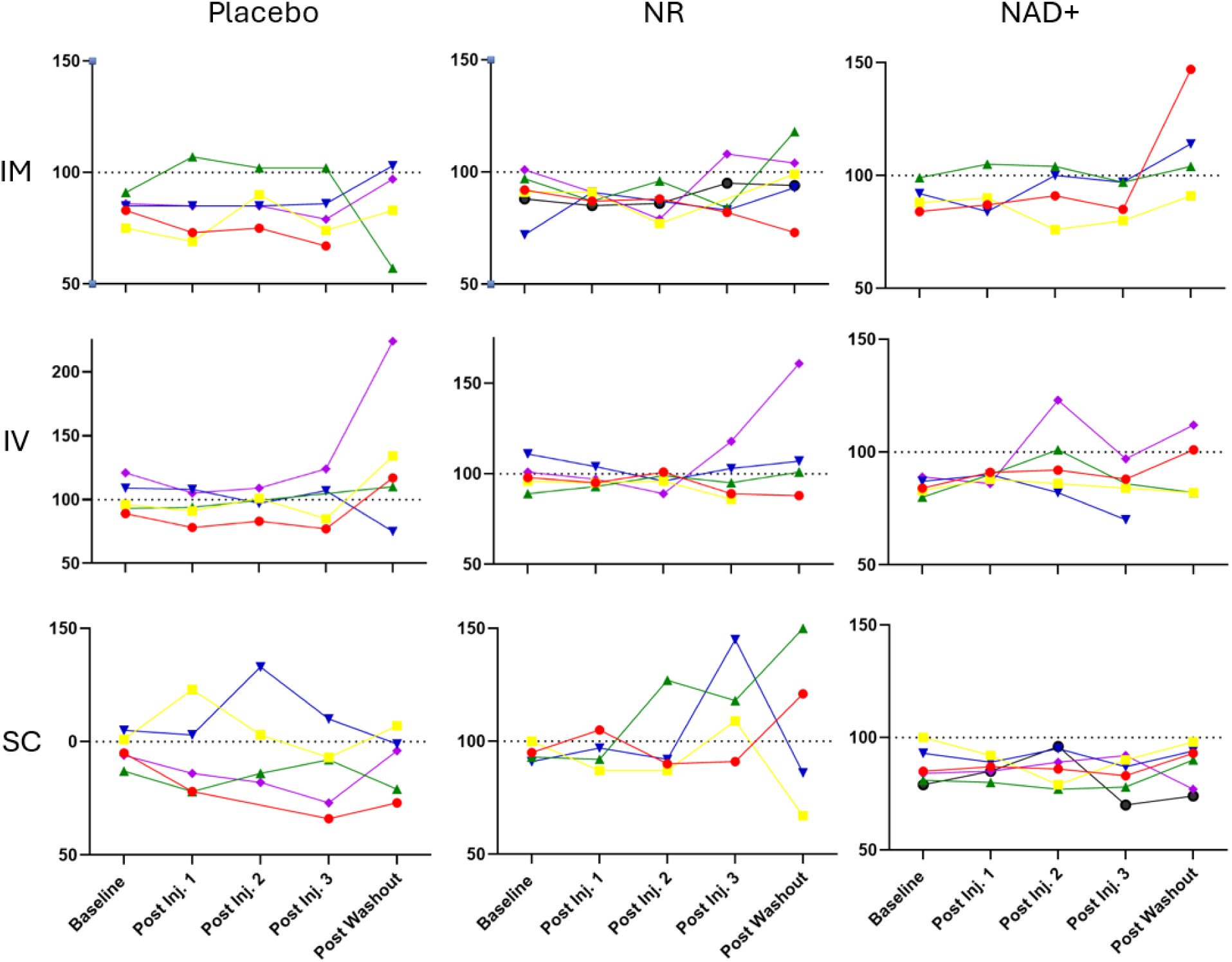
Trial 1, Individual Participant Changes in Fasted Blood Glucose. Fasted blood glucose (mg/dL) was measured on days 1 at baseline, and 90 minutes after the injection on days 1, 2, and 3. Additionally, fasted blood glucose was measured on day 10, following the 7-day washout period. Normal blood glucose is considered <100 mg/dL, which is indicated as the dashed line on the figures. Each line represents measurements in individual study participants in each group. Colors across graphs, thus, do not correspond to the same person. The glucose responses for each participant were variable and did not follow a specific pattern.

**Figure 4.**
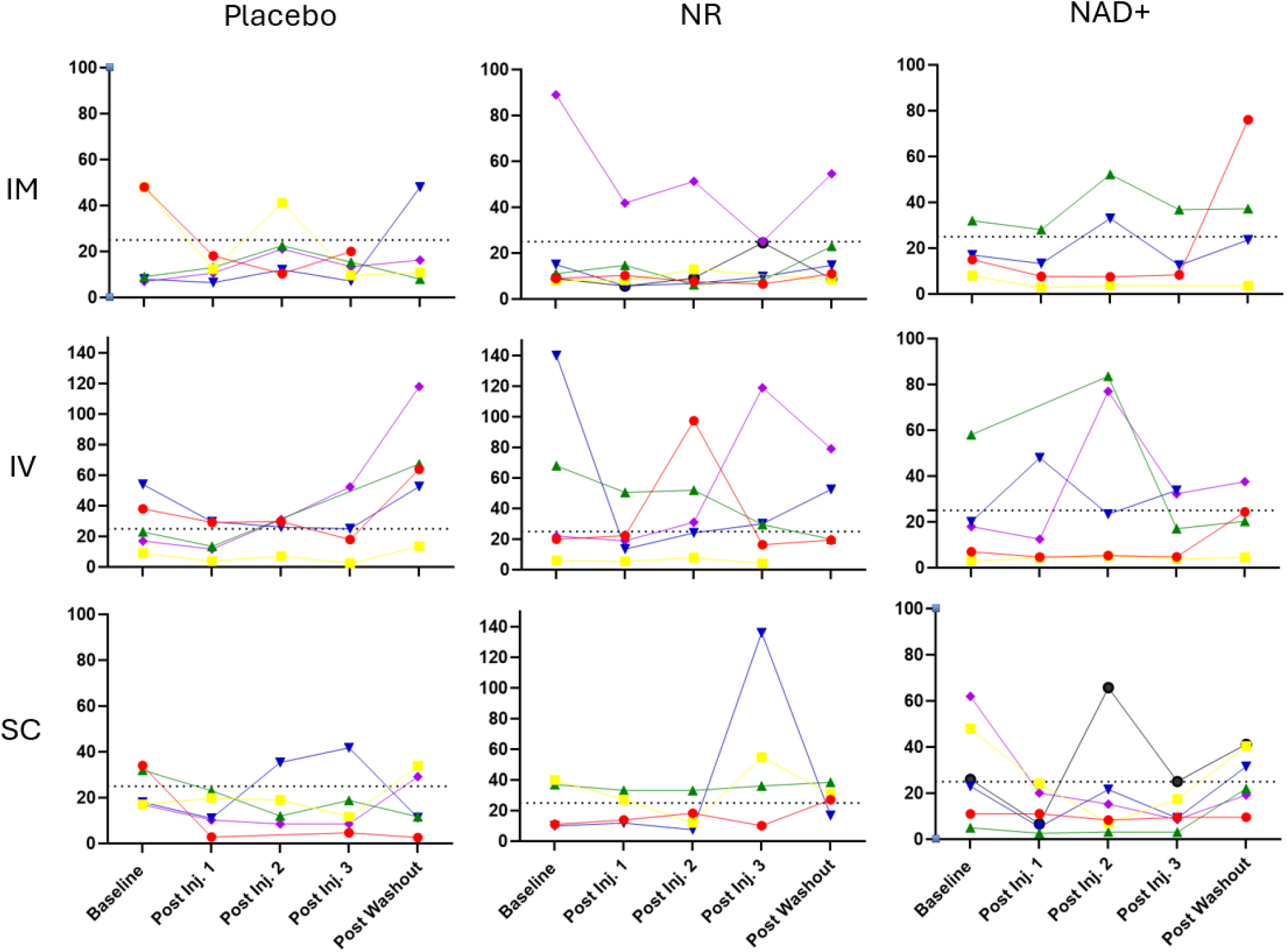
Trial 1, Individual Changes in Fasted Serum Insulin. Fasted serum insulin (μIU/ml) measured on days 1 at baseline, and 90 minutes after the injection on days 1, 2, and 3. Additionally, fasted serum insulin was measured on day 10, following the 7-day washout period. Normal fasted serum insulin is considered <25 μIU/ml, which is indicated as the dashed line on the figures. Each line represents measurements in individual study participants in each group. Colors across graphs, thus, do not correspond to the same person. The insulin responses for each participant were variable and did not follow a specific pattern.

Inflammatory markers of high-sensitivity C-reactive protein (hsCRP), serum viscosity and erythrocyte sedimentation rate (ESR) were measured at baseline, 90 minutes after each injection and after the washout (Figure 5). Levels of hsCRP stayed within the normal range of <0.3mg/L, though levels in the NR IV and NAD+ IV groups followed different patterns than the placebo group, post-washout. This does not appear to be a treatment effect for NR nor NAD+ but rather reflects an isolated increase in inflammation in one of the placebo group participants. Subcutaneous NR participants had elevated hsCRP at baseline, compared to the NAD+ and placebo SC groups. The second and third injections of NR reduced hsCRP in this group, which were further reduced following the washout. Serum viscosity was not different between groups. Baseline ESR appeared elevated when comparing - the NR and placebo groups for SC administration, and after one injection, differences between the groups were not observed, potentially demonstrating a positive effect of the single injection of NR.

**Figure 5.**
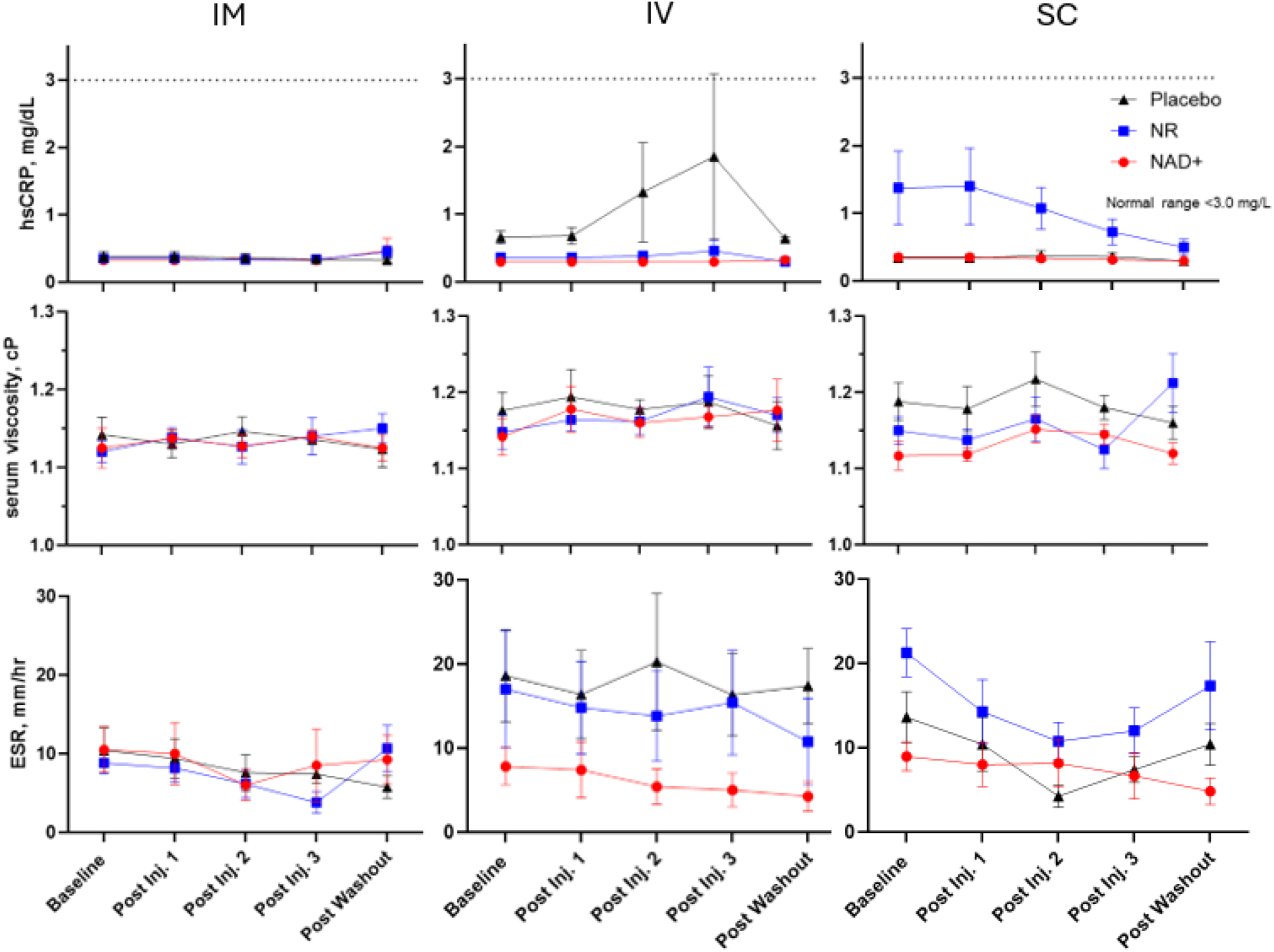
Trial 1 Blood Inflammatory Markers. High-sensitivity C-reactive protein (hsCRP), serum viscosity, and erythrocyte sedimentation rate (ESR) were measured in study participants as clinical indicators of inflammation. Values are presented as the group mean ± SEM,.

#### 3.1.5. Trial 1 Adverse Events & Subjective Injection Experience

Both NR and NAD+ injections were well-tolerated, as no participants withdrew from discomfort or other injection-related concerns, nor were any reported severe or unexpected adverse events attributable to the test materials. One adverse event was reported during Trial 1 (NAD+ group). This event was classified as moderate in severity and not related to the intervention by the PI and medical monitor. The reported adverse event was a Mycoplasma pneumoniae infection developed by one participant after direct contact with a contagious person at their place of employment, and was recovered at the time of the visit.

At the time of and a few minutes following each injection, the participants were asked to describe their experience with the injection. The responses were open-ended and not standardized. These responses were summarized and categorized, and the frequency of the observations for all three injections are reported in Tables 4 and 5. The sum of the frequency of negative reported experiences is listed on each table. Pain and discomfort were the most consistent and frequently reported adverse experiences across interventions and routes of administration. Of the nine interventions, NR IM appeared to be the most painful at the time of the injection. Other than pain and discomfort, participant responses were unique by test material and route of administration.

### 3.2. Trial 2

#### 3.2.1. Trial 2 Participants

A total of 67 individuals living in or near Adventura, Florida were screened and assessed for eligibility, and 40 participants were selected for randomization. One individual was excluded from receiving a dose due to hypertension at baseline. The 39 remaining participants were randomized to 4 arms: 50mg of NR via IM injection, 50mg of NR via SC injection, 100mg of NR via IM injection, 100mg of NR via SC injection. The participants each received 3 injections, 1 each day at clinic visits 1-3, and returned on Day 10 for post-washout assessments and to initiate Phase 2, the 2-arm, at-home portion of the study. All participants who received 50 mg NR, regardless of route of administration, were placed into the 50 mg SC for Phase 2. Similarly, all participants receiving a 100 mg dose, regardless of route of administration were placed in the 100 mg SC group. All 39 participants completed all site visits, however, two participants in phase 2 (1 from each group) were determined to have had significant deviation from the protocol and were not included in per protocol calculations (Figure 6).

**Figure 6.**
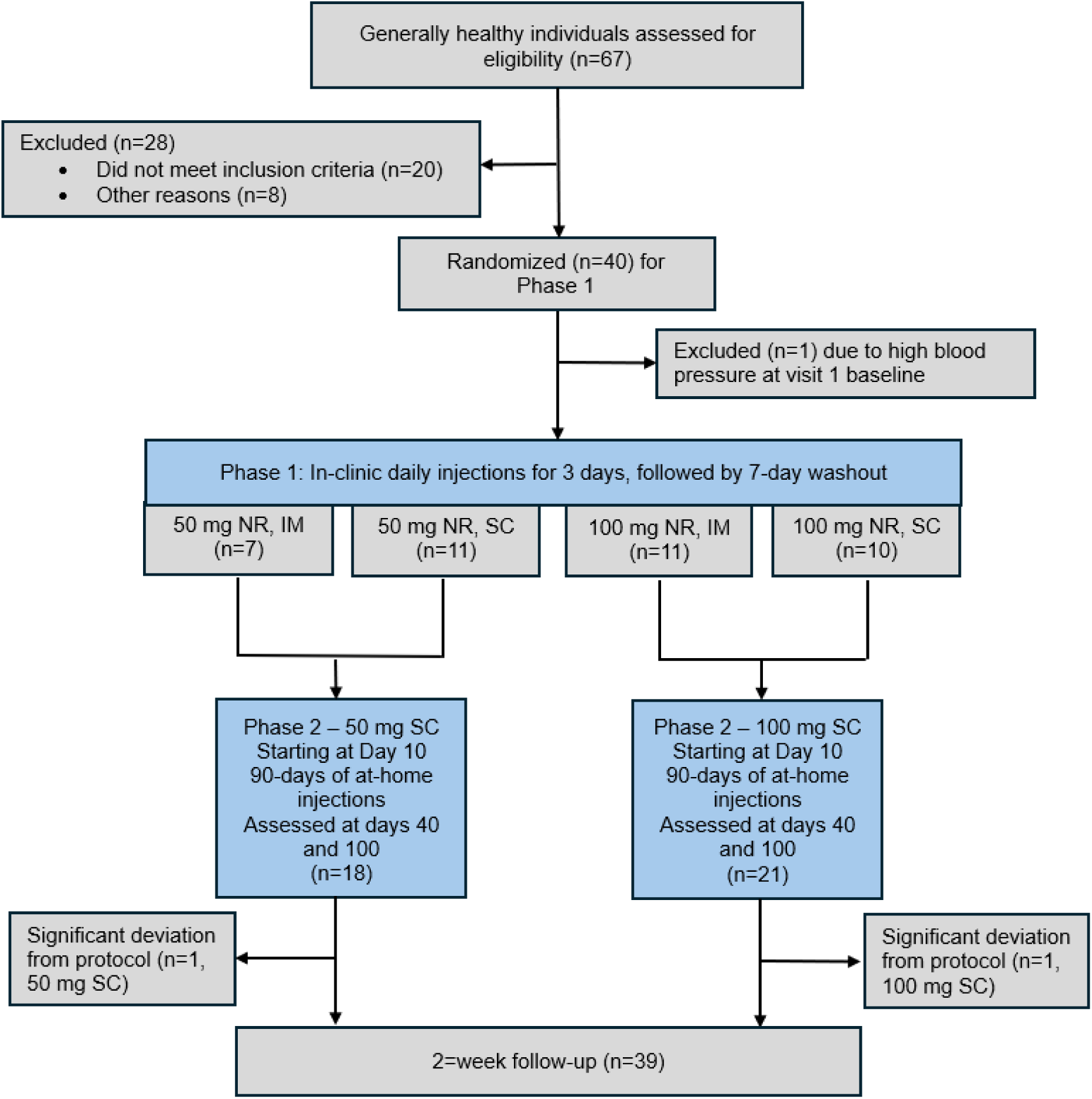
Trial 2 Consort Flow Diagram.

##### Trial 2 Trial Demographics

In Trial 2, 59% of the participants were female, however randomization did not incorporate a gender balance, thus the 50 mg IM and 50 mg SC groups skew female and while the 100 mg IM group has more males. The 100 mg SC group is balanced for males and females.

The average age of the participants was 46.7 (±1.9 SEM) and 46% of the participants self-reported as White, non-Hispanic. All participants had Fatigue Assessment scores ≥22, indicating some degree of more than normal fatigue as determined by the Fatigue Assessment Scale (FAS). For the entire study population, the minimum, maximum, and median FAS were 22, 45, and 28, respectively.

#### 3.2.2. Trial 2 Vitals

Participant vitals were assessed across Phase 1 and Phase 2 of the trial. In Phase 1, baseline vitals, along with measurements at the 90-minute and 180-minute time points were assessed on visit (V) 1 (V1). On visit 2 (V2) and visit 3 (V3), vitals were collected 90 minutes post-injection. Subsequent measurements were taken once on visit 4 (V4), visit 5 (V5), and visit 6 (V6). Participants in the 50 mg NR SC injection cohort experienced a statistically significant difference in SBP between baseline (130.70±6.14) and 90-minute (120.10±4.74) time points. There was also a significant difference in heart rate (HR) within the three time points (70.00±1.93, 67.82±1.41, 80.27±4.00) on V1 for the 100 mg IM injection group.

Significant changes were observed for RR in the 50mg IM group across the 3 time points on V1 and V1-V6 overall. Further Tukey pairwise analysis showed this difference was present specifically when comparing V1 and V6. A significant difference for peripheral capillary oxygen saturation (SpO_2_) was observed in the 100mg SC group during V1 between baseline and 90-minute time point, as well as when comparing all three time points. Lastly, upon measuring body temperature, the 50 mg SC group displayed a statistically significant ANOVA; however, pairwise comparisons of V1 and V4 or V1 and V6 did not reach significance.

#### 3.2.4 Trial 2 Blood Markers

A total of 22 blood biomarkers were collected in this study (Table 7): albumin, alkaline phosphatase, alanine aminotransferase (ALT), aspartate aminotransferase (AST), bilirubin total (Bili total), blood urea nitrogen (BUN), calcium, carbon dioxide, chloride, creatinine, glucose, hematocrit, hemoglobin, mean corpuscular hemoglobin (MCH), mean corpuscular hemoglobin concentration (MCHC), mean corpuscular volume (MCV), platelet count, potassium, protein total, red blood cell (RBC) count, sodium, white blood cell (WBC) count. While there were statistically significant changes in some biomarkers, in general, changes observed were within normal ranges and did not constitute a safety signal. Such conclusions were confirmed by a third-party clinician.

**Table 6.** Trial 2 Baseline Demographics

| Intent to Treat (ITT), N=39 |  |  |  |  |  |
| --- | --- | --- | --- | --- | --- |
| Demographic | Injection Administration Group | 50 mg IM | 100 mg IM | 50 mg SC | 100 mg SC |
|  | N | 7 | 11 | 11 | 10 |
| Sex | Female, n | 5 (71%) | 4 (36%) | 9 (82%) | 5 (50%) |
|  | Male, n | 2 (29%) | 7 (64%) | 2 (18%) | 5 (50%) |
| Age | Age, years ± SEM | 44.6 ± 2.65 | 47.2 ± 2.74 | 46.9 ± 3.95 | 47.4 ± 5.22 |
| Self-Described Race/Ethnicity | Asian, n | 1 | 1 | 1 | 0 |
|  | Black & Asian | 0 | 0 | 0 | 1 |
|  | Brazilian | 0 | 1 | 0 | 0 |
|  | East Indian | 0 | 0 | 1 | 0 |
|  | Hispanic | 3 | 1 | 2 | 4 |
|  | Latin | 0 | 1 | 0 | 0 |
|  | Mixed Ethnicity (ME) | 0 | 2 | 0 | 1 |
|  | Other | 0 | 0 | 1 | 0 |
|  | White | 3 | 4 | 6 | 4 |
|  | White/ME | 0 | 1 | 0 | 0 |
| BMI, kg/m<sup>2</sup> ± SEM | | 25.1 ± 2.00 | 26.9 ± 1.48 | 24.0 ± 0.93 | 28.1 ± 2.28 |
| Smoker, n (%) |  | 0 (0%) | 1 (9.1%) | 1 (9.1%) | 1 (10%) |
| Illicit Drug Use, n (%) |  | 0 (0%) | 1 (9.1%) | 0 (0%) | 0 (0%) |
| Alcohol use | None, n (%) | 3 (42.9%) | 6 (54.5%) | 6 (54.5%) | 4 (40%) |
|  | 1-3x/wk, n (%) | 2 (28.6%) | 2 (18.2%) | 3 (27.3%) | 3 (30%) |
|  | 4-7x/wk, n (%) | 1 (14.3%) | 3 (27.3%) | 1 (9.1%) | 4 (40%) |
|  | No answer, n (%) | 1 (14.3%) | 0 (0%) | 1 (9.1%) | 0 (0%) |
| Average Screening FAS ± SEM | | 27.6 ± 1.32 | 26.5 ± 1.22 | 28.2 ± 1.85 | 30.8 ± 2.55 |

**Table 7.**
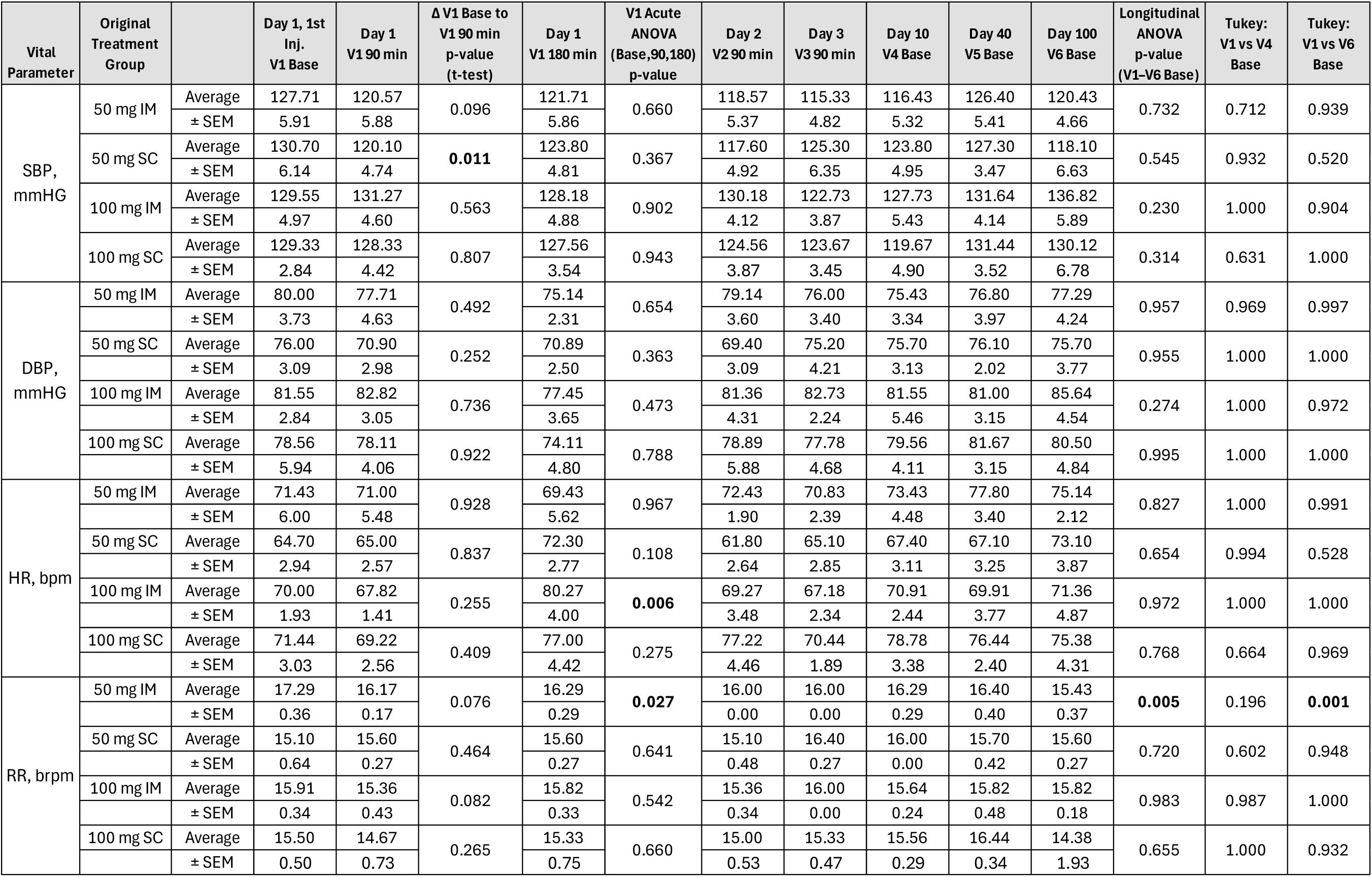
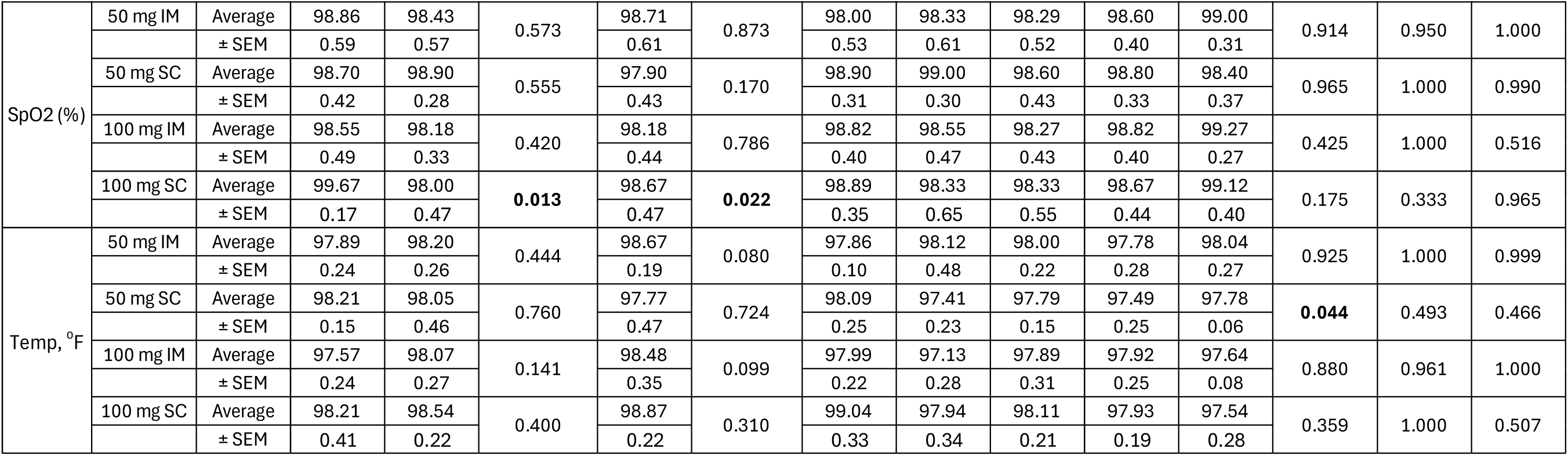
Trial 2 Vitals.

| Vital Parameter | Original Treatment Group | | Day 1, 1st Inj. V1 Base | Day 1 V1 90 min | $\Delta$ V1 Base to V1 90 min p-value (t-test) | Day 1 V1 180 min | V1 Acute ANOVA (Base,90,180) p-value | Day 2 V2 90 min | Day 3 V3 90 min | Day 10 V4 Base | Day 40 V5 Base | Day 100 V6 Base | Longitudinal ANOVA p-value (V1-V6 Base) | Tukey: V1 vs V4 Base | Tukey: V1 vs V6 Base |
| --- | --- | --- | --- | --- | --- | --- | --- | --- | --- | --- | --- | --- | --- | --- | --- |
| SBP, mmHG | 50 mg IM | Average | 127.71 | 120.57 | 0.096 | 121.71 | 0.660 | 118.57 | 115.33 | 116.43 | 126.40 | 120.43 | 0.732 | 0.712 | 0.939 |
|  |  | ± SEM | 5.91 | 5.88 |  | 5.86 |  | 5.37 | 4.82 | 5.32 | 5.41 | 4.66 |  |  |  |
|  | 50 mg SC | Average | 130.70 | 120.10 | **0.011** | 123.80 | 0.367 | 117.60 | 125.30 | 123.80 | 127.30 | 118.10 | 0.545 | 0.932 | 0.520 |
|  |  | ± SEM | 6.14 | 4.74 |  | 4.81 |  | 4.92 | 6.35 | 4.95 | 3.47 | 6.63 |  |  |  |
|  | 100 mg IM | Average | 129.55 | 131.27 | 0.563 | 128.18 | 0.902 | 130.18 | 122.73 | 127.73 | 131.64 | 136.82 | 0.230 | 1.000 | 0.904 |
|  |  | ± SEM | 4.97 | 4.60 |  | 4.88 |  | 4.12 | 3.87 | 5.43 | 4.14 | 5.89 |  |  |  |
|  | 100 mg SC | Average | 129.33 | 128.33 | 0.807 | 127.56 | 0.943 | 124.56 | 123.67 | 119.67 | 131.44 | 130.12 | 0.314 | 0.631 | 1.000 |
|  |  | ± SEM | 2.84 | 4.42 |  | 3.54 |  | 3.87 | 3.45 | 4.90 | 3.52 | 6.78 |  |  |  |
| DBP, mmHG | 50 mg IM | Average | 80.00 | 77.71 | 0.492 | 75.14 | 0.654 | 79.14 | 76.00 | 75.43 | 76.80 | 77.29 | 0.957 | 0.969 | 0.997 |
|  |  | ± SEM | 3.73 | 4.63 |  | 2.31 |  | 3.60 | 3.40 | 3.34 | 3.97 | 4.24 |  |  |  |
|  | 50 mg SC | Average | 76.00 | 70.90 | 0.252 | 70.89 | 0.363 | 69.40 | 75.20 | 75.70 | 76.10 | 75.70 | 0.955 | 1.000 | 1.000 |
|  |  | ± SEM | 3.09 | 2.98 |  | 2.50 |  | 3.09 | 4.21 | 3.13 | 2.02 | 3.77 |  |  |  |
|  | 100 mg IM | Average | 81.55 | 82.82 | 0.736 | 77.45 | 0.473 | 81.36 | 82.73 | 81.55 | 81.00 | 85.64 | 0.274 | 1.000 | 0.972 |
|  |  | ± SEM | 2.84 | 3.05 |  | 3.65 |  | 4.31 | 2.24 | 5.46 | 3.15 | 4.54 |  |  |  |
|  | 100 mg SC | Average | 78.56 | 78.11 | 0.922 | 74.11 | 0.788 | 78.89 | 77.78 | 79.56 | 81.67 | 80.50 | 0.995 | 1.000 | 1.000 |
|  |  | ± SEM | 5.94 | 4.06 |  | 4.80 |  | 5.88 | 4.68 | 4.11 | 3.15 | 4.84 |  |  |  |
| HR, bpm | 50 mg IM | Average | 71.43 | 71.00 | 0.928 | 69.43 | 0.967 | 72.43 | 70.83 | 73.43 | 77.80 | 75.14 | 0.827 | 1.000 | 0.991 |
|  |  | ± SEM | 6.00 | 5.48 |  | 5.62 |  | 1.90 | 2.39 | 4.48 | 3.40 | 2.12 |  |  |  |
|  | 50 mg SC | Average | 64.70 | 65.00 | 0.837 | 72.30 | 0.108 | 61.80 | 65.10 | 67.40 | 67.10 | 73.10 | 0.654 | 0.994 | 0.528 |
|  |  | ± SEM | 2.94 | 2.57 |  | 2.77 |  | 2.64 | 2.85 | 3.11 | 3.25 | 3.87 |  |  |  |
|  | 100 mg IM | Average | 70.00 | 67.82 | 0.255 | 80.27 | **0.006** | 69.27 | 67.18 | 70.91 | 69.91 | 71.36 | 0.972 | 1.000 | 1.000 |
|  |  | ± SEM | 1.93 | 1.41 |  | 4.00 |  | 3.48 | 2.34 | 2.44 | 3.77 | 4.87 |  |  |  |
|  | 100 mg SC | Average | 71.44 | 69.22 | 0.409 | 77.00 | 0.275 | 77.22 | 70.44 | 78.78 | 76.44 | 75.38 | 0.768 | 0.664 | 0.969 |
|  |  | ± SEM | 3.03 | 2.56 |  | 4.42 |  | 4.46 | 1.89 | 3.38 | 2.40 | 4.31 |  |  |  |
| RR, brpm | 50 mg IM | Average | 17.29 | 16.17 | 0.076 | 16.29 | **0.027** | 16.00 | 16.00 | 16.29 | 16.40 | 15.43 | **0.005** | 0.196 | **0.001** |
|  |  | ± SEM | 0.36 | 0.17 |  | 0.29 |  | 0.00 | 0.00 | 0.29 | 0.40 | 0.37 |  |  |  |
|  | 50 mg SC | Average | 15.10 | 15.60 | 0.464 | 15.60 | 0.641 | 15.10 | 16.40 | 16.00 | 15.70 | 15.60 | 0.720 | 0.602 | 0.948 |
|  |  | ± SEM | 0.64 | 0.27 |  | 0.27 |  | 0.48 | 0.27 | 0.00 | 0.42 | 0.27 |  |  |  |
|  | 100 mg IM | Average | 15.91 | 15.36 | 0.082 | 15.82 | 0.542 | 15.36 | 16.00 | 15.64 | 15.82 | 15.82 | 0.983 | 0.987 | 1.000 |
|  |  | ± SEM | 0.34 | 0.43 |  | 0.33 |  | 0.34 | 0.00 | 0.24 | 0.48 | 0.18 |  |  |  |
|  | 100 mg SC | Average | 15.50 | 14.67 | 0.265 | 15.33 | 0.660 | 15.00 | 15.33 | 15.56 | 16.44 | 14.38 | 0.655 | 1.000 | 0.932 |
|  |  | ± SEM | 0.50 | 0.73 |  | 0.75 |  | 0.53 | 0.47 | 0.29 | 0.34 | 1.93 |  |  |  |

Table 7. continued
| Vital Parameter | Original Treatment Group | | Day 1, 1st Inj. V1 Base | Day 1 V1 90 min | $\Delta$ V1 Base to V1 90 min p-value (t-test) | Day 1 V1 180 min | V1 Acute ANOVA (Base,90,180) p-value | Day 2 V2 90 min | Day 3 V3 90 min | Day 10 V4 Base | Day 40 V5 Base | Day 100 V6 Base | Longitudinal ANOVA p-value (V1-V6 Base) | Tukey: V1 vs V4 Base | Tukey: V1 vs V6 Base |
| --- | --- | --- | --- | --- | --- | --- | --- | --- | --- | --- | --- | --- | --- | --- | --- |
| SpO2 (%) | 50 mg IM | Average | 98.86 | 98.43 | 0.573 | 98.71 | 0.873 | 98.00 | 98.33 | 98.29 | 98.60 | 99.00 | 0.914 | 0.950 | 1.000 |
| | | ± SEM | 0.59 | 0.57 | | 0.61 | | 0.53 | 0.61 | 0.52 | 0.40 | 0.31 | | | |
|  | 50 mg SC | Average | 98.70 | 98.90 | 0.555 | 97.90 | 0.170 | 98.90 | 99.00 | 98.60 | 98.80 | 98.40 | 0.965 | 1.000 | 0.990 |
| | | ± SEM | 0.42 | 0.28 | | 0.43 | | 0.31 | 0.30 | 0.43 | 0.33 | 0.37 | | | |
|  | 100 mg IM | Average | 98.55 | 98.18 | 0.420 | 98.18 | 0.786 | 98.82 | 98.55 | 98.27 | 98.82 | 99.27 | 0.425 | 1.000 | 0.516 |
| | | ± SEM | 0.49 | 0.33 | | 0.44 | | 0.40 | 0.47 | 0.43 | 0.40 | 0.27 | | | |
|  | 100 mg SC | Average | 99.67 | 98.00 | **0.013** | 98.67 | **0.022** | 98.89 | 98.33 | 98.33 | 98.67 | 99.12 | 0.175 | 0.333 | 0.965 |
| | | ± SEM | 0.17 | 0.47 | | 0.47 | | 0.35 | 0.65 | 0.55 | 0.44 | 0.40 | | | |
| Temp, °F | 50 mg IM | Average | 97.89 | 98.20 | 0.444 | 98.67 | 0.080 | 97.86 | 98.12 | 98.00 | 97.78 | 98.04 | 0.925 | 1.000 | 0.999 |
| | | ± SEM | 0.24 | 0.26 | | 0.19 | | 0.10 | 0.48 | 0.22 | 0.28 | 0.27 | | | |
|  | 50 mg SC | Average | 98.21 | 98.05 | 0.760 | 97.77 | 0.724 | 98.09 | 97.41 | 97.79 | 97.49 | 97.78 | **0.044** | 0.493 | 0.466 |
| | | ± SEM | 0.15 | 0.46 | | 0.47 | | 0.25 | 0.23 | 0.15 | 0.25 | 0.06 | | | |
|  | 100 mg IM | Average | 97.57 | 98.07 | 0.141 | 98.48 | 0.099 | 97.99 | 97.13 | 97.89 | 97.92 | 97.64 | 0.880 | 0.961 | 1.000 |
| | | ± SEM | 0.24 | 0.27 | | 0.35 | | 0.22 | 0.28 | 0.31 | 0.25 | 0.08 | | | |
|  | 100 mg SC | Average | 98.21 | 98.54 | 0.400 | 98.87 | 0.310 | 99.04 | 97.94 | 98.11 | 97.93 | 97.54 | 0.359 | 1.000 | 0.507 |
| | | ± SEM | 0.41 | 0.22 | | 0.22 | | 0.33 | 0.34 | 0.21 | 0.19 | 0.28 | | | |

A significant change for albumin was observed between V1 baseline (BL) and V4 for the 50 mg IM, 50 mg SC, and 100 mg IM groups, p=0.039, 0.040, 0.006, respectively. BUN had a significant change between V1 BL and V1 post-injection in 50 mg IM and SC groups. There was a change in the carbon dioxide levels between V1 BL and V4 for the 50 mg SC group (p=0.048), and a change from V1 BL to V6 in the 100 mg SC group (p=0.009). When comparing chloride levels from V1 BL to V6, both IM groups (50 mg and 100 mg) experienced a significant change, with the 100 mg IM group also noting a change between V1 BL and V4. The only change noted for the biomarker creatinine was in the 100mg SC group from V1 BL to V4, p=0.003.

Both 50 mg groups and the 100mg IM group experienced significant decreases in blood glucose concentrations between V1 BL and V1 post-injection, as depicted in Table 8. Figure 7B shows a sustained elevation of blood glucose concentrations of an individual participant in the 100 mg SC group exceeding the normal range at baseline and throughout the timepoints assessed. Interestingly, hematocrit, hemoglobin, and RBC levels experienced the same pattern of significant change between V1 BL measurement and V4 in the 50 mg SC and 100 mg IM participant groups. MCHC did have a few notable changes, namely,. significant increase between V1 BL and V6 for the 50 mg IM, 50 mg SC, and 100 mg SC groups, with the 100 mg SC group also demonstrating an increase between V1 BL and V1 post-injection. As for MCV, like MCHC, both the 50 mg IM and 50 mg SC participant groups demonstrated a statistically significant difference between V1 BL and V6.

**Figure 7.**
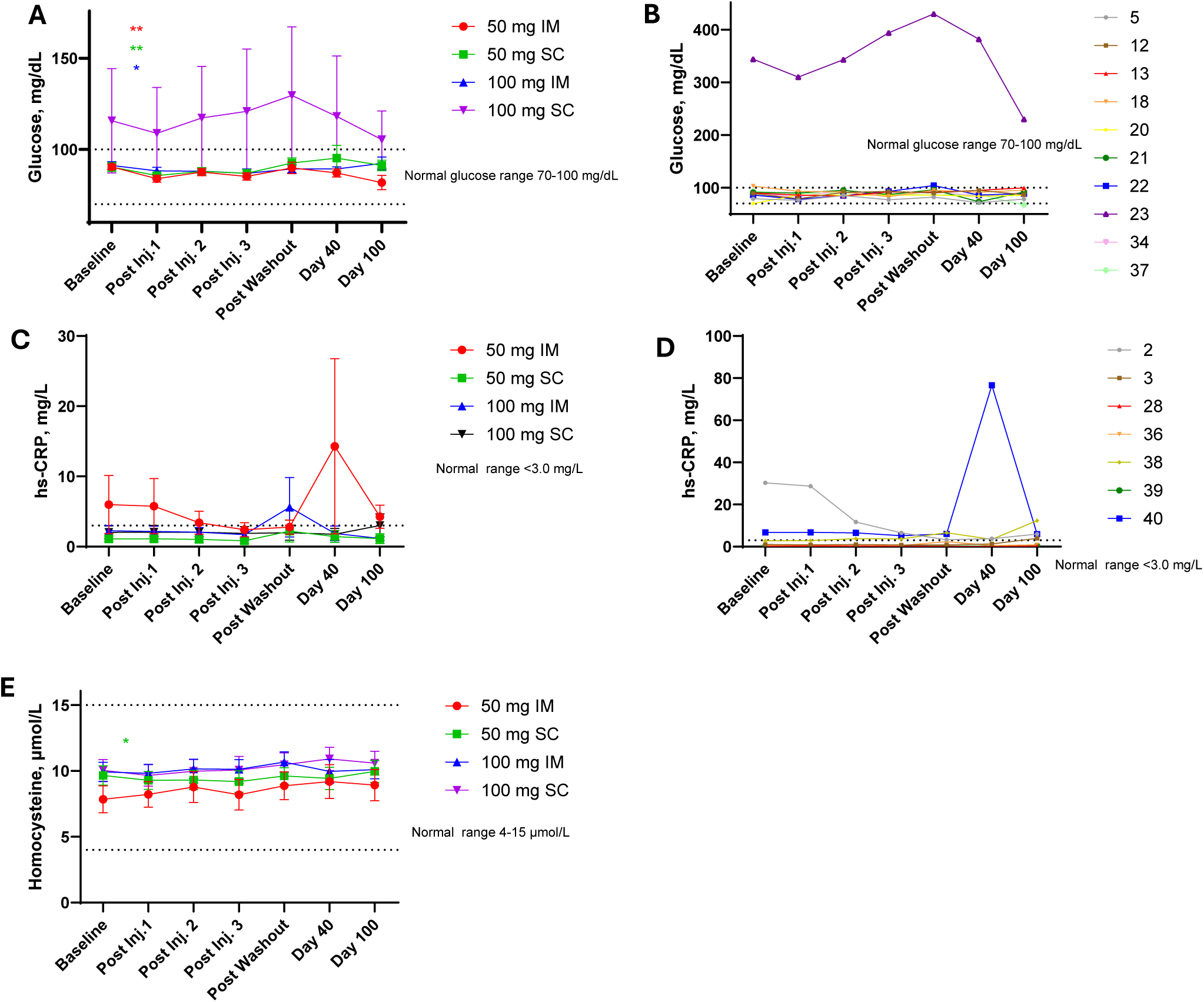
Trial 2 Glucose, hs-CRP, and homocysteine. **A**. Serum glucose by phase 1 treatment group. Following the first injection, 50 mg IM, 50 mg SC, and 100 mg IM resulted in a statistically significant decrease in serum glucose. **B**. Individual participant serum glucose in the phase 1, 100 mg SC group, demonstrating one participant as an outlier. **C**. The hs-CRP by phase 1 treatment group. **D**. Individual participant hs-CRP in the phase 1, 50 mg IM group. **E**. Mean homocysteine ±SEM by original treatment group. Following the first injection, 50 mg SC resulted in a statistically significant decrease in homocysteine. Values in A, C, and E are represented as the mean ±SEM, *p<0.05, **p<0.01, ***p<0.001.

**Table 8.** Trial 2 Blood Biomarkers. Mean ±SEM. Bold indicates values outside of normal reference ranges.

| Blood Biomarker | Phase 1 Treatment Group | V1 Day 1 Baseline | V1 Day 1 Post Inj. 1 | V2 Day 2 Post Inj. 2 | V3 Day 3 Post Inj. 3 | V4 Day 10 Post Washout | V5 Day 40 | V6 Day 100 | V1 BL → Post Inj. 1 stat. sig. & change | V1 BL→V4 stat. sig & change | V1 BL→V6 stat. sig & change |
| --- | --- | --- | --- | --- | --- | --- | --- | --- | --- | --- | --- |
| **Albumin** | 50 mg IM | 4.42±0.11 | 4.45±0.09 | 4.53±0.10 | 4.46±0.09 | 4.55±0.10 | 4.51±0.10 | 4.33±0.14 | ns | p=0.039<br>↑ (+2.7%) | ns |
|  | 50 mg SC | 4.57±0.14 | 4.46±0.09 | 4.52±0.11 | 4.44±0.12 | 4.31±0.11 | 4.40±0.11 | 4.40±0.11 | ns | p=0.040<br>↓ (-5.7%) | ns |
|  | 100 mg IM | 4.54±0.08 | 4.51±0.09 | 4.51±0.08 | 4.53±0.06 | 4.34±0.09 | 4.43±0.08 | 4.45±0.08 | ns | p=0.006<br>↓ (-4.5%) | ns |
|  | 100 mg SC | 4.40±0.08 | 4.47±0.10 | 4.44±0.13 | 4.53±0.14 | 4.50±0.14 | 4.61±0.13 | 4.47±0.15 | ns | ns | ns |
| **Alkaline Phosphatase** | 50 mg IM | 72.59±7.78 | 72.81±7.38 | 73.94±7.99 | 71.97±7.98 | 74.47±8.36 | 72.70±9.83 | 73.67±8.66 | ns | ns | ns |
|  | 50 mg SC | 57.96±7.05 | 57.87±7.50 | 56.97±7.18 | 55.67±6.94 | 57.46±6.80 | 56.80±6.28 | 55.33±6.17 | ns | ns | ns |
|  | 100 mg IM | 63.41±5.88 | 62.06±5.92 | 63.91±6.27 | 63.81±6.63 | 61.95±6.81 | 58.92±3.20 | 62.46±5.20 | ns | ns | ns |
|  | 100 mg SC | 66.57±3.79 | 66.48±3.74 | 66.49±2.59 | 66.80±3.00 | 70.33±3.61 | 69.98±2.50 | 69.41±4.06 | ns | ns | ns |
| **ALT** | 50 mg IM | 29.63±9.25 | 28.84±8.85 | 28.19±9.14 | 27.10±9.49 | 23.49±5.98 | 20.05±4.56 | 24.77±4.29 | ns | ns | ns |
|  | 50 mg SC | 27.77±7.23 | 27.09±7.22 | 25.85±6.52 | 23.41±4.64 | 25.34±4.08 | 20.48±2.86 | 21.35±2.52 | ns | ns | ns |
|  | 100 mg IM | 34.99±5.33 | 34.58±5.36 | 37.15±6.96 | 36.16±6.59 | 32.73±5.17 | 28.40±2.47 | 37.78±7.70 | ns | ns | ns |
|  | 100 mg SC | **42.84±18.10** | **42.60±18.13** | **42.06±17.93** | **41.82±17.39** | **50.76±26.10** | **49.48±22.54** | 39.31±8.67 | ns | ns | ns |
| **AST** | 50 mg IM | 24.77±3.59 | 23.94±3.28 | 23.49±3.86 | 22.93±3.86 | 22.37±2.72 | 22.48±2.78 | 22.44±2.27 | ns | ns | ns |
|  | 50 mg SC | 26.27±4.29 | 25.70±4.60 | 23.11±3.60 | 21.87±1.83 | 29.21±9.11 | 22.32±1.95 | 21.71±2.10 | ns | ns | ns |
|  | 100 mg IM | 30.36±3.73 | 30.28±3.81 | 31.41±4.25 | 29.81±4.03 | 26.65±2.67 | 29.45±3.36 | 32.36±5.07 | ns | ns | ns |
|  | 100 mg SC | 31.76±9.73 | 31.58±9.62 | 30.59±9.27 | 29.92±8.39 | 35.84±14.74 | 35.13±10.93 | 27.43±4.06 | ns | ns | ns |
| **Bili Total** | 50 mg IM | 0.47±0.05 | 0.52±0.04 | 0.47±0.06 | 0.51±0.03 | 0.46±0.05 | 0.49±0.06 | 0.47±0.06 | ns | ns | ns |
|  | 50 mg SC | 0.54±0.05 | 0.55±0.04 | 0.45±0.04 | 0.53±0.04 | 0.43±0.04 | 0.32±0.05 | 0.46±0.04 | ns | ns | ns |
|  | 100 mg IM | 0.54±0.09 | 0.58±0.08 | 0.59±0.10 | 0.60±0.10 | 0.53±0.09 | 0.63±0.10 | 0.61±0.08 | ns | ns | ns |
|  | 100 mg SC | 0.61±0.07 | 0.63±0.07 | 0.55±0.07 | 0.55±0.06 | 0.59±0.05 | 0.65±0.07 | 0.67±0.05 | ns | ns | ns |
| **BUN** | 50 mg IM | 12.87±1.26 | 12.06±1.29 | 14.69±2.04 | 12.77±2.18 | 13.70±1.42 | 12.84±1.92 | 14.66±1.58 | p<0.001<br>↓ (-6.3%) | ns | ns |
|  | 50 mg SC | 12.88±1.49 | 12.21±1.48 | 13.64±0.98 | 13.16±1.18 | 13.10±1.37 | 15.88±1.83 | 14.36±1.18 | p=0.019<br>↓ (-5.2%) | ns | ns |
|  | 100 mg IM | 16.18±1.99 | 14.38±1.54 | 14.98±2.02 | 14.39±1.82 | 15.23±1.52 | 12.84±0.79 | 15.03±1.31 | ns | ns | ns |
|  | 100 mg SC | 13.22±2.23 | 12.77±2.15 | 12.83±1.60 | 12.34±1.48 | 13.49±1.49 | 13.42±1.56 | 15.16±1.92 | ns | ns | ns |
| Calcium | 50 mg IM | 9.37±0.12 | 9.50±0.09 | 9.71±0.12 | 9.51±0.10 | 9.45±0.09 | 9.47±0.13 | 9.48±0.13 | ns | ns | ns |
|  | 50 mg SC | 9.56±0.10 | 9.46±0.09 | 9.48±0.12 | 9.45±0.13 | 9.41±0.13 | 9.31±0.14 | 9.56±0.09 | ns | ns | ns |
|  | 100 mg IM | 9.52±0.09 | 9.52±0.13 | 9.58±0.11 | 9.58±0.10 | 9.42±0.09 | 9.43±0.06 | 9.50±0.09 | ns | ns | ns |
|  | 100 mg SC | 9.43±0.09 | 9.47±0.08 | 9.54±0.14 | 9.57±0.11 | 9.59±0.15 | 9.65±0.11 | 9.64±0.15 | ns | ns | ns |
| Carbon Dioxide | 50 mg IM | 21.59±0.45 | 21.87±0.62 | 21.99±0.62 | 22.56±0.44 | 22.21±0.36 | 21.33±0.66 | 21.66±0.72 | ns | ns | ns |
|  | 50 mg SC | 22.19±0.59 | 21.62±0.73 | 22.96±0.81 | 22.20±0.76 | 23.17±0.50 | 22.01±0.53 | 22.54±0.45 | ns | p=0.048<br>↑ (+4.4%) | ns |
|  | 100 mg IM | 22.47±0.75 | 23.15±0.49 | 23.13±0.63 | 22.95±0.53 | 22.58±0.61 | 22.61±0.58 | 23.15±0.73 | ns | ns | ns |
|  | 100 mg SC | 20.94±0.26 | 21.54±0.65 | 22.70±0.46 | 22.61±0.33 | 21.69±0.61 | 21.79±0.53 | 23.30±0.68 | ns | ns | p=0.009<br>↑ (+11.2%) |
| Chloride | 50 mg IM | 103.54±0.62 | 103.01±0.68 | 102.23±0.40 | 102.71±0.86 | 103.84±0.55 | 102.67±1.15 | 102.00±0.50 | ns | ns | p=0.010<br>↓ (-1.5%) |
|  | 50 mg SC | 103.07±0.66 | 103.48±0.38 | 103.66±0.49 | 103.70±0.51 | 104.26±0.35 | 104.38±0.25 | 102.93±0.50 | ns | ns | ns |
|  | 100 mg IM | 103.01±0.60 | 102.68±0.61 | 103.56±0.67 | 103.41±0.50 | 103.95±0.65 | 104.02±0.57 | 102.05±0.52 | ns | p=0.013<br>↑ (+0.9%) | p=0.015<br>↓ (-0.9%) |
|  | 100 mg SC | 102.32±0.88 | 102.38±0.77 | 102.61±1.08 | 103.09±0.99 | 103.00±0.94 | 102.81±0.95 | 101.20±0.74 | ns | ns | ns |
| Creatinine | 50 mg IM | 0.77±0.08 | 0.76±0.08 | 0.78±0.08 | 0.75±0.08 | 0.76±0.08 | 0.78±0.08 | 0.74±0.07 | ns | ns | ns |
|  | 50 mg SC | 0.80±0.07 | 0.78±0.06 | 0.78±0.06 | 0.78±0.06 | 0.76±0.05 | 0.76±0.06 | 0.76±0.05 | ns | ns | ns |
|  | 100 mg IM | **0.97±0.08** | **0.95±0.08** | **0.94±0.08** | **0.94±0.07** | **0.95±0.07** | **0.98±0.08** | **0.91±0.06** | ns | ns | ns |
|  | 100 mg SC | **0.94±0.08** | **0.95±0.07** | **0.93±0.07** | **0.95±0.07** | **0.99±0.08** | **0.95±0.08** | **0.96±0.07** | ns | p=0.003<br>↑ (+5.4%) | ns |
| Glucose | 50 mg IM | 90.47±1.39 | 83.96±1.90 | 87.56±2.04 | 85.21±2.00 | 89.91±3.19 | 87.08±2.23 | 81.83±3.97 | p=0.003<br>↓ (-7.2%) | ns | ns |
|  | 50 mg SC | 90.41±2.47 | 85.71±1.48 | 87.96±2.10 | 86.90±2.14 | 92.53±2.80 | 95.24±7.03 | 91.23±2.90 | p=0.009<br>↓ (-5.2%) | ns | ns |
|  | 100 mg IM | 91.25±2.01 | 88.25±1.93 | 88.05±1.54 | 86.94±2.41 | 89.35±2.13 | 89.29±1.71 | 92.35±3.44 | p=0.041<br>↓ (-3.3%) | ns | ns |
|  | 100 mg SC | **115.76±28.68** | **108.86±25.21** | **117.28±28.25** | **120.91±34.18** | **129.62±37.61** | **118.11±33.10** | **105.37±15.71** | ns | ns | ns |
| Hematocrit | 50 mg IM | 40.83±1.50 | 41.29±0.67 | 42.07±1.26 | 41.26±0.99 | 41.59±1.09 | 42.08±1.51 | 40.59±0.78 | ns | ns | ns |
|  | 50 mg SC | 40.99±0.79 | 40.73±0.82 | 40.16±0.70 | 40.27±0.32 | **39.74±0.74** | **39.22±0.75** | **39.47±0.52** | ns | p=0.047<br>↓ (-3.0%) | ns |
|  | 100 mg IM | 43.95±1.52 | 43.45±1.50 | 43.20±1.49 | 43.93±1.67 | 42.46±1.58 | 42.28±1.86 | 42.72±1.77 | ns | p=0.026<br>↓ (-3.4%) | ns |
|  | 100 mg SC | 41.77±1.10 | 41.71±1.15 | 41.57±1.15 | 41.39±0.89 | 41.96±1.01 | 41.53±1.02 | 41.66±1.05 | ns | ns | ns |
| Hemoglobin | 50 mg IM | **13.41±0.46** | 13.73±0.28 | 14.07±0.48 | 13.80±0.37 | 13.73±0.41 | 14.08±0.49 | 13.74±0.36 | ns | ns | ns |
|  | 50 mg SC | **13.32±0.29** | **13.26±0.28** | **13.21±0.29** | **13.19±0.22** | **12.96±0.28** | **12.90±0.29** | **13.09±0.23** | ns | p=0.047<br>↓ (-2.7%) | ns |
|  | 100 mg IM | 14.62±0.57 | 14.50±0.57 | 14.46±0.56 | 14.50±0.57 | 14.08±0.57 | 14.07±0.70 | 14.33±0.65 | ns | p=0.002<br>↓ (-3.7%) | ns |
|  | 100 mg SC | 13.77±0.37 | 13.90±0.36 | 13.88±0.38 | **13.69±0.37** | 13.91±0.34 | 13.92±0.33 | 13.90±0.41 | ns | ns | ns |
| MCH | 50 mg IM | 30.83±0.47 | 30.91±0.42 | 30.76±0.45 | 30.47±0.52 | 30.61±0.63 | 30.93±0.75 | 30.61±0.71 | ns | ns | ns |
|  | 50 mg SC | 29.56±0.54 | 29.63±0.45 | 29.73±0.54 | 29.67±0.48 | 29.61±0.46 | 29.96±0.57 | 29.75±0.69 | ns | ns | ns |
|  | 100 mg IM | 30.02±0.59 | 29.93±0.58 | 29.93±0.58 | 29.85±0.62 | 29.85±0.58 | 30.14±0.65 | 29.85±0.69 | ns | ns | ns |
|  | 100 mg SC | 30.66±0.66 | 30.89±0.69 | 30.78±0.68 | 30.31±0.64 | 30.66±0.62 | 30.82±0.67 | 30.54±0.66 | ns | ns | ns |
| MCHC | 50 mg IM | 32.87±0.51 | 33.23±0.34 | 33.43±0.46 | 33.46±0.31 | 33.00±0.32 | 33.48±0.37 | 33.84±0.46 | ns | ns | p=0.010<br>↑ (+3.0%) |
|  | 50 mg SC | 32.48±0.25 | 32.55±0.24 | 32.87±0.30 | 32.73±0.33 | 32.61±0.31 | 32.88±0.26 | 33.15±0.33 | ns | ns | p=0.001<br>↑ (+2.1%) |
|  | 100 mg IM | 33.21±0.22 | 33.33±0.28 | 33.45±0.27 | 32.99±0.25 | 33.12±0.28 | 33.19±0.29 | 33.47±0.26 | ns | ns | ns |
|  | 100 mg SC | 32.97±0.38 | 33.36±0.40 | 33.41±0.33 | 33.06±0.33 | 33.16±0.33 | 33.56±0.39 | 33.37±0.45 | p=0.032<br>↑ (+1.2%) | ns | p=0.024<br>↑ (+1.2%) |
| MCV | 50 mg IM | **93.74±0.70** | **93.01±0.85** | 91.96±0.94 | 91.16±1.30 | **92.67±1.49** | **92.35±1.62** | 90.44±1.29 | ns | ns | p=0.019<br>↓ (-3.5%) |
|  | 50 mg SC | 90.89±1.27 | 90.98±1.16 | 90.36±1.12 | 90.63±1.13 | 90.84±1.14 | 91.14±1.34 | 89.67±1.47 | ns | ns | p=0.042<br>↓ (-1.3%) |
|  | 100 mg IM | 90.40±1.64 | 89.83±1.62 | 89.48±1.57 | 90.43±1.84 | 90.05±1.55 | 90.75±1.56 | 89.17±1.89 | ns | ns | ns |
|  | 100 mg SC | **92.91±1.51** | **92.57±1.43** | 92.07±1.39 | 91.74±1.62 | **92.39±1.58** | 91.83±1.35 | 91.52±1.24 | ns | ns | ns |
| Platelet Count | 50 mg IM | 278.43±38.73 | 282.71±28.67 | 296.14±31.97 | 295.14±29.61 | 321.43±48.16 | 279.33±33.27 | 299.86±37.19 | ns | ns | ns |
|  | 50 mg SC | 298.00±25.86 | 285.50±24.13 | 286.70±24.88 | 284.30±22.49 | 283.30±21.86 | 261.10±25.04 | 273.20±24.40 | ns | ns | ns |
|  | 100 mg IM | 253.36±15.65 | 254.00±14.16 | 257.64±16.87 | 259.27±17.65 | 268.82±18.53 | 270.45±18.64 | 271.09±16.22 | ns | ns | p=0.046<br>↑ (+7.0%) |
|  | 100 mg SC | 282.44±20.62 | 279.44±19.96 | 290.89±21.42 | 288.56±24.33 | 286.44±24.59 | 282.56±24.99 | 236.67±33.99 | ns | ns | ns |
| Potassium | 50 mg IM | 4.35±0.11 | 4.29±0.14 | 4.40±0.19 | 4.24±0.15 | 4.43±0.14 | 4.30±0.15 | 4.25±0.14 | ns | ns | ns |
|  | 50 mg SC | 4.32±0.06 | 4.32±0.09 | 4.45±0.14 | 4.33±0.12 | 4.33±0.08 | 4.27±0.09 | 4.28±0.08 | ns | ns | ns |
|  | 100 mg IM | 4.39±0.10 | 4.37±0.09 | 4.43±0.08 | 4.34±0.11 | 4.35±0.08 | 4.38±0.11 | 4.32±0.10 | ns | ns | ns |
|  | 100 mg SC | 4.49±0.10 | 4.39±0.08 | 4.42±0.10 | 4.50±0.08 | 4.56±0.10 | 4.56±0.09 | 4.60±0.11 | ns | ns | ns |
| Protein Total | 50 mg IM | 7.10±0.24 | 7.12±0.19 | 7.33±0.25 | 7.15±0.24 | 7.12±0.27 | 7.25±0.30 | 7.03±0.27 | ns | ns | ns |
|  | 50 mg SC | 7.23±0.16 | 7.05±0.11 | 7.01±0.08 | 6.89±0.12 | 6.88±0.11 | 6.82±0.12 | 6.98±0.09 | ns | ns | ns |
|  | 100 mg IM | 7.08±0.07 | 7.14±0.09 | 7.20±0.12 | 7.16±0.08 | 6.87±0.13 | 6.92±0.11 | 7.01±0.12 | ns | ns | ns |
|  | 100 mg SC | 6.78±0.08 | 6.88±0.12 | 6.87±0.16 | 6.97±0.14 | 7.00±0.18 | 7.09±0.16 | 6.99±0.15 | ns | ns | ns |
| RBC | 50 mg IM | 4.36±0.18 | 4.44±0.10 | 4.58±0.15 | 4.53±0.12 | 4.50±0.16 | 4.57±0.22 | 4.50±0.14 | ns | ns | ns |
|  | 50 mg SC | 4.51±0.08 | 4.48±0.09 | 4.45±0.09 | 4.45±0.07 | 4.38±0.09 | 4.31±0.09 | 4.41±0.10 | ns | p=0.029<br>↓ (-3.0%) | ns |
|  | 100 mg IM | 4.88±0.18 | 4.85±0.17 | 4.84±0.17 | 4.87±0.19 | 4.73±0.20 | 4.67±0.22 | 4.81±0.21 | ns | p=0.020<br>↓ (-2.9%) | ns |
|  | 100 mg SC | 4.51±0.17 | 4.52±0.18 | 4.53±0.17 | 4.53±0.16 | 4.56±0.17 | 4.54±0.17 | 4.57±0.17 | ns | ns | ns |
| Sodium | 50 mg IM | 139.03±0.58 | 138.77±0.63 | 138.59±0.28 | 139.26±0.52 | 139.79±1.01 | 138.03±0.64 | 138.86±0.71 | ns | ns | ns |
|  | 50 mg SC | 138.98±0.63 | 139.16±0.53 | 139.14±0.66 | 139.13±0.44 | 140.39±0.42 | 139.41±0.67 | 139.83±0.57 | ns | p=0.027<br>↑ (+1.0%) | ns |
|  | 100 mg IM | 139.62±0.58 | 139.14±0.55 | 140.36±0.62 | 139.84±0.54 | 140.45±0.77 | 140.26±0.85 | 139.26±0.62 | ns | ns | ns |
|  | 100 mg SC | 137.63±0.57 | 138.26±0.67 | 138.89±0.77 | 139.36±0.84 | 138.72±0.83 | 138.76±0.71 | 139.03±0.56 | p=0.030<br>↑ (+0.5%) | ns | p=0.039<br>↑ (+1.0%) |
| WBC | 50 mg IM | 5.78±0.33 | 6.52±0.49 | 6.24±0.44 | 6.56±0.46 | 6.59±0.70 | 6.60±0.63 | 6.87±0.71 | p=0.041<br>↑ (+12.8%) | ns | ns |
|  | 50 mg SC | 5.46±0.53 | 5.61±0.48 | 5.60±0.42 | 5.59±0.39 | 5.53±0.43 | 5.30±0.34 | 5.63±0.58 | ns | ns | ns |
|  | 100 mg IM | 5.94±0.55 | 5.99±0.50 | 6.24±0.63 | 5.63±0.35 | 5.31±0.38 | 5.62±0.65 | 5.32±0.47 | ns | ns | ns |
|  | 100 mg SC | 5.52±0.37 | 5.64±0.41 | 5.65±0.38 | 5.91±0.52 | 5.94±0.60 | 5.71±0.37 | 6.39±0.78 | ns | ns | ns |

Upon measuring platelet count, the only significant change was confined to the 100 mg IM group across V1 BL to V6 timepoints. Notable changes were observed for sodium levels in both SC groups. A significant increase was observed in the 50mg SC group from V1 BL to V4 measurements, whereas a significant increase was observed in the 100mg SC group between V1 BL and V1 post-injection and V1 BL to V6 measurements. The 50 mg SC cohort also demonstrated a significant decrease in homocysteine levels following the first injection (Figure 7E). Lastly, there was only one statistically significant difference for WBC count in the 50mg IM group between V1 BL and V1 post-injection.

#### 3.2.5 Trial 2 Participant Reported Experience, Injection Site Reactions and Adverse Events

A summary of participant described experiences in Trial 2 are provided in Table 9. Whie NR injections were generally well tolerated, 45.9% of participants across groups reported pain more than 2 minutes after the injection and 43.2% reported muscle soreness or tightness. Redness and itching occurred following SC injections in 52.6% and 57.9% of participants, respectively. Reported increase in alertness or energy ranged from 10% in the 50 mg SC group to 71.4% for the 50 mg IM group, whereas tiredness was reported by 24.3% of participants. Notably, the 100 mg IM cohort had 0% incidence of redness, swelling, itching, or bruising after their injection, and less than 50% incidence rate of feeling pain over 2 minutes after injection, warmth and muscle soreness. Interestingly, flushing/heat sensation was only reported with SC injections.

**Table 9.** Trial 2 Frequency of participant reported experiences following in-clinic injections (Phase 1)

| Participant reported injection experience | 50 mg IM % | 50 mg SC % | 100 mg IM % | 100 mg SC % | 50 mg % | 100 mg % | IM % | SC % | Pooled % |
| --- | --- | --- | --- | --- | --- | --- | --- | --- | --- |
| Bruising | 0.0 | 0.0 | 0.0 | 11.1 | 0.0 | 5.0 | 0.0 | 5.3 | 2.7 |
| Decrease in energy | 0.0 | 0.0 | 18.2 | 11.1 | 0.0 | 15.0 | 11.1 | 5.3 | 8.1 |
| Dizziness/ light-headed | 14.3 | 0.0 | 0.0 | 11.1 | 5.9 | 5.0 | 5.6 | 5.3 | 5.4 |
| Flushing/heat sensation | 0.0 | 10.0 | 0.0 | 22.2 | 5.9 | 10.0 | 0.0 | 15.8 | 8.1 |
| Headache | 14.3 | 10.0 | 0.0 | 22.2 | 11.8 | 10.0 | 5.6 | 15.8 | 10.8 |
| Increase in alertness/energy | 71.4 | 10.0 | 18.2 | 11.1 | 35.3 | 15.0 | 38.9 | 10.5 | 24.3 |
| Itching | 0.0 | 60.0 | 0.0 | 55.6 | 35.3 | 25.0 | 0.0 | 57.9 | 29.7 |
| Muscle soreness/tightness | 85.7 | 20.0 | 45.5 | 33.3 | 47.1 | 40.0 | 61.1 | 26.3 | 43.2 |
| Pain > 2min after injection | 71.4 | 40.0 | 18.2 | 66.7 | 52.9 | 40.0 | 38.9 | 52.6 | 45.9 |
| Redness | 14.3 | 50.0 | 0.0 | 55.6 | 35.3 | 25.0 | 5.6 | 52.6 | 29.7 |
| Swelling | 14.3 | 0.0 | 0.0 | 22.2 | 5.9 | 10.0 | 5.6 | 10.5 | 8.1 |
| Tiredness/sleepy | 28.6 | 20.0 | 27.3 | 22.2 | 23.5 | 25.0 | 27.8 | 21.1 | 24.3 |
| Warmth | 28.6 | 30.0 | 18.2 | 11.1 | 29.4 | 15.0 | 22.2 | 21.1 | 21.6 |

During Trial 2, seven adverse events (AE) were reported, with only one AE reported during the at-home phase of the trial (Table 10). None of the adverse events were deemed serious by the PI, though a significant pain experience did occur with one participant following what was likely an injection into a superficial nerve. At the 14-day follow-up (Day 114, two weeks after the at-home injections stopped), 7 participants reported AEs experienced after the completion of the study (cold/flu, food poisoning, and gastrointestinal virus), though none of the AEs were deemed to be related to the test material (data not shown)

**Table 10.** Trial 2 Reported Adverse Events During the Trial

| Route | Dose, mg | Visit | AE Description | Severity | Serious | Related to IP | Expected | Action Taken / Details | Clinician Notes |
| --- | --- | --- | --- | --- | --- | --- | --- | --- | --- |
| SC | 055 | Visit 1 | Fever 100.6°F | Mild | No | Unknown | No | None | N/A |
| SC | 100 | Visit 1 | Headache 2/10; slightly dizzy | Mild | No | Unknown | No | None | N/A |
| IM | 050 | Visit 3 | Fever after injection | Mild | No | Unknown | No | None | No signs or symptoms |
| IM | 050 | Visit 2 | Temp 100.6°F at 90-min check | Mild | No | Unknown | No | Discharged | N/A |
| SC | 100 | At-home | Sharp injection-site pain with radiating pain on a dermatome path; no swelling; improved with hot compress and Tylenol; no pain the next morning. | Important Medical | No | Unknown | No | Hot compress x1 hr; Tylenol; injection noted to be shallow angle/slow (~60 sec for 1 mL) | Likely hit superficial nerve due to shallow angle of needle and slow injection. |
| SC | 100 | Visit 4 | Critical Lab Value - Glucose 430 | Mild | No | No | No | Critical lab value received from Vibrant lab on 12/5/25. | Clinician made contact with participant to discuss lab value. Participant was advised to follow-up with PCP |
| IM | 100 | Visit 6 | Hypertension - 176/116 | Mild | No | No | No | Reported to PI and advised participant to follow up with PCP. | N/A |

## 4. Discussion

The present report describes the safety findings from two pilot clinical trials evaluating the parenteral administration of pharmaceutical-grade NR (Niagen^®^ Plus) via IV, IM, and SC routes. In Trial 1, all three routes were assessed and compared to NAD+ administered via equivalent routes, whereas Trial 2 was limited to NR via IM and SC routes, without a placebo control. Across both trials, NR administered parenterally was generally well-tolerated, with no treatment-related serious adverse events. Though there were isolated, minor changes in health and safety parameters, the alterations were generally not clinically meaningful and displayed no evidence of dose-dependent effects.

The comparative post-hoc data from Trial 1 are suggestive of a tolerability pattern that varies as a function of route of administration. For IV bolus administration, NR generated fewer total subjective adverse experiences than NAD+, both during the infusion period, and in the post-infusion window. These findings corroborate previous clinical findings^33,59^, and are consistent with the mechanistic premise that IV NAD+ increases extracellular NAD+, which in turn stimulates an immune signaling cascade that induces proinflammatory responses, which could manifest as pain and muscle tightness ^31,38^. On the other hand, for SC administration, both NR and NAD+ performed comparably immediately upon and post injection. In the IM administration conditions, NAD+ appeared better tolerated than NR, with fewer intra-injection complaints with NAD+ injections, and the same number of complaints post-injection for both compounds. The physiological basis explaining these route-dependent differences are unclear and are somewhat at odds with short-term animal data demonstrating superior tolerability following NR administered via IM over SC and IV^60^.

In Trial 2, adverse experience frequency did not appear dose-responsive for IM injections of NR, whereas for SC injections flushing/heat sensation, headache, and pain were more frequently reported with the higher dose. As this study is pilot in design, only one person reporting the experience for the lower dose and two participants for the higher dose is not sufficient to draw conclusions of a dose response. Important to note are the methodologic limitations that strongly limit the interpretability of these subjective experience observations – symptom data were collected via open-ended, unprompted self-report that were entirely qualitative in nature, rather than a standardized, validated instrument. Thus, this introduces heterogeneity in how participants described their experiences and raises the likelihood that the apparent differences in adverse event frequencies may represent reporting style differences, rather than true pharmacologic differences. Further accentuating these limitations is the absence of formal severity grading. As such, it is therefore not possible to determine whether a report of “pain/discomfort” represented a brief, transient sensation or a more prolonged experience, nor to what extent the participant was affected. Therefore, frequency counts without severity context limits the clinical utility and interpretability of these data. Therefore, a more standardized approach evaluating participant experience for IM and SC injections warrants investigation in larger study populations before conclusions can be made.

In Trial 1, fasted blood glucose responses were highly variable, with some participants displaying increases and others displaying decreases, following injections. No significant differences were detected across NR, NAD+, or placebo groups for any route of administration. Variability in glucose changes were noted in both active and placebo arms, suggesting biological variation rather than a pharmacologic effect of either NAD+ or NR intervention. On the other hand, decreases in fasting blood glucose concentrations were documented in Trial 2. While the magnitude of change was quantitatively modest and values remained within normal reference ranges, the directional consistency across the three groups (50 mg IM, 50 mg SC, and 100 mg IM) may possibly signal an acute insulin-sensitizing or glucose flux-modulating effect of NR. These results are somewhat corroborated by Reyna et al., who identified very small reductions in glycosylated hemoglobin (HbA1C) values among a cohort of NR-IV users after a 30-day follow-up period^38^. Nevertheless, the discordant findings between Trials 1 and 2 warrant caution in overinterpretation. In contrast, the 100 mg SC group did not demonstrate post-injection changes in blood glucose concentrations. An investigation of the individual-level data revealed that this null finding may be attributable to a single participant with markedly elevated glucose levels at baseline, i.e., >400 mg/dL likely secondary to unrecognized, uncontrolled hyperglycemia, throughout the study period. These levels were significantly above normal, healthy ranges and were well beyond the values observed in all other participants, likely influencing group mean and variance. However, no formal sensitivity analyses were conducted to quantify the influence of this observation, and thus its impact on group-level outcomes remain uncertain. The participant was retained in all primary analyses; no post-hoc exclusion or adjustments were performed. Though an active diabetes diagnosis was grounds for exclusion in both trials, it may be prudent for future trials to employ both fasting plasma glucose and glycosylated hemoglobin (HbA1c) values to exclude participants with unrecognized uncontrolled glycemic dysregulation.

Oral administration of NR has resulted in modified reductions in SBP, that have not been validated in larger population studies^61^. Changes in vital signs across both trials were generally modest and lacked consistent directional or dose-dependent patterns. The most notable finding was an observed reduction in SBP of roughly 12 mmHg in the NR IV group, following the 7-day washout period, in Trial 1. A parallel acute SBP reduction of approximately 10 mmHg was also documented in the 50 mg SC group at 90 mins post injection, in Trial 2. Though the extent of the reduction may appear quantitatively impressive, the changes were not sustained longitudinally in Trial 2, in which there were no significant group-level SBP differences at completion vs. baseline. The effects were also inconsistent across groups, further complicating the interpretation of the isolated changes. Therefore, the physiological or clinical significance of these findings are unclear. Of note, longer-term oral supplementation with NAD+ precursor compounds have previously been demonstrated to exert anti-hypertensive effects, in some, but not all clinical studies ^62–65^.

Further studies, with adequate statistical power, are required to determine the durability of parenteral NR’s effects on vascular health, and whether these effects vary based on baseline health characteristics.

Though a significant change in HR was observed in the 100 mg IM group at the 180-min mark on Day 1 in Trial 2, there were no sustained HR changes observed at other visits. As such, this is likely explained by biological variation or other measurement-related effects (e.g., postural differences), rather than an effect of NR. Similarly, significant differences were observed in RR and SpO_2_ at certain timepoints; however, the extent of the changes were minor and within normal physiological ranges. Additionally, the changes in were not clinically meaningful and difficult to attributable to NR. The modest changes in HR, RR, and SpO_2_ do not appear to be treatment related nor clinically relevant.

In clinical trials of orally administered NR, reductions in inflammatory markers, particularly proinflammatory cytokines, appears to be consistent, and is the most replicated benefit of NR supplementation.^48,49,66–71^ Chronic inflammation, appears to drive NAD+ decline, and may be associated with the aging phenotype ^72^. Here, inflammatory markers were incorporated as blood safety markers to determine if injections resulted in an immune response. There was a modest anticipation that individuals with overweight or obesity would have elevated baseline inflammation^73,74^ that would respond to NR and/or NAD+. In trial 1, subcutaneous NR resulted in observed reductions in inflammation, when compared to the placebo, driven by the elevated hsCRP at baseline in this group, which was not replicated in the NAD+ and placebo groups. Neither NR nor NAD+ appeared to induce a systemic inflammatory reaction in response to the injections. However, potential therapeutic benefits of NR or NAD+ injections in individuals with elevated baseline inflammation may be warranted. Further placebo-controlled studies are needed in individuals with elevated baseline inflammation to validate these findings. In trial 2, an impact of NR on inflammation was not observed. Additionally, incorporation of the analysis of inflammatory cytokines in addition to hsCRP, serum viscosity, and ESR would further strengthen the findings related to inflammation augmentation.

There are several key limitations to the present two studies that warrant acknowledgement. Importantly, both were exploratory pilot trials; therefore, the absence of statistically significant adverse safety findings should be interpreted as a preliminary safety signal, not a definitive characterization of the long-term safety profile of injectable NR. The modest sample size precludes the ability to detect rare adverse events, or conclusively demonstrate differences in safety by relevant subgroups, e.g., age, sex, bodyweight.

The absence of a placebo arm in Trial 2 limits causal inferences regarding injectable NR; indeed, some of the observed changes may have arisen due to confounders, such as background lifestyle factors, random variation, temporal effects, or regression to the mean. Without a placebo, every within-subject paired t-test is uninterpretable as a treatment effect. The intervention effect cannot be distinguished from time-of-day, fasting state, regression to the mean, or repeated venipuncture artifact. All reported within-arm changes from baseline should be considered descriptive only, and support hypothesis generation.

Values that were determined to be statistically significant for SBP and homocysteine are statistical only, not causal, and require validation in a larger, powered study.

The open-label nature of Trial 2 also introduces the potential for expectancy bias that may influence participant-reported outcomes. In both trials, a formal standardized adverse event grading instrument such as the Common Terminology Criteria for Adverse Events (CTCAE) was not employed, which limits the ability to make severity-graded comparisons across groups. This is consistent with the exploratory pilot design of both studies. In Trial 1, active structured AE monitoring was conducted at seven protocol-specified timepoints per injection day, with severity and causality assessed per a six-category and seven-category classification framework, respectively, in accordance with 21 CFR 312.32. Open-ended participant responses were collected as a supplementary measure and are analyzed separately as described in Section 2.1.9. In Trial 2, participant-reported symptom data were also collected without formal severity grading. Although these data collectively show that the injections are tolerable and do not result in safety signals, future research assessing participant experience using systematic and validated scales will better capture and quantify the participant experience.

Unfortunately, neither trial incorporated complete blood count data, which would provide further evidence of safety. Variability in blood markers of glucose, inflammation, homocysteine, and hsCRP suggest individual and unique responses to the injections, compromising general safety conclusions. Larger, powered studies, utilizing similar study designs, but employing full comprehensive metabolic panels and complete blood counts will help to develop population safety data, and reveal treatment-related changes in safety markers, if they exist. Additionally, preclinical, repeat-dose toxicology studies are needed to reveal similarities and variations between routes of NR administration and target organs of toxicity, which are currently unknown.

Lastly, both of these studies incorporated secondary and exploratory efficacy outcomes that are not reported in this manuscript. Future reporting that addresses these findings may further elucidate the mechanisms and benefits of NR injections.

1. 5. Conclusion

Two pilot clinical trials explored safety outcomes related to NR injections. The results suggest that NR injections, while associated with some discomfort, were generally well-tolerated and not associated with safety risks in this limited sample. Further, home administration was feasible under the present protocol-defined conditions. These preliminary data suggest possible acute effects on systolic blood pressure and inflammatory markers, which warrant confirmation in future research studies.

## Statement of Contribution

- All authors were required to review and provide comments on the manuscript prior to submission.
- YNE facilitated project management and study initiation for both clinical trials. She was involved in the development of the study protocols, data analysis and interpretation, data visualization, manuscript writing, editing, and final manuscript review.
- JK contributed to data analysis and visualization, manuscript writing, editing, and final manuscript review.
- SS served as the lead statistician for Trial 2, and contributed to data analysis and interpretation, manuscript editing and final manuscript review.
- RI conducted data analysis and interpretation, data visualization, manuscript writing and editing, and final manuscript review.
- JM contributed to manuscript writing and editing, and final manuscript review.
- AR served as the clinical study manager for Trial 2, was responsible for final protocol development, clinical trial management, data collection, data analysis and interpretation, and final review of the manuscript.
- JC served as the contracted PI for Trial 1, was responsible for obtaining IRB approval, final study protocol, supervising the trial, data collection, data analysis and interpretation, and final review of the manuscript.
- LH contributed to Trial 1 on project methodology, data curation, study investigation, and resource management.
- KF contributed to Trial 1 on data curation, study investigation, resource management, and validation.
- JJ contributed to Trial 1 to project methodology, data curation, study investigation, and supervision.
- AS was involved in study ideation and the development of study protocols for both trials, manuscript editing, and final manuscript review.

## Declaration of Generative AI Use

- Generative AI was not used in the development of this manuscript

## Data Availability

All data produced in the present study are available upon reasonable request to the corresponding author.

## Acknowledgements

Thank you to the study participants for both trials.

- Thank you to Jessie Hawkins-Cavanaugh, PhD and Halland Chen, MD for serving as the study PIs for Trial 1 and Trial 2, respectively. Your contributions to the study concept, IRB approval, clinical trial management, data collection, data analysis and data interpretation were extremely valuable and we appreciate your efforts. We are looking forward to co-authoring future manuscripts on these topics.
- Thank you to the staff and clinicians at Impact Health and Nutraceutical Research Institute for your contributions, with special acknowledgement to Tracy Allen, Harry Pindell, Sofia Gadelov, Courtney Pindell, Emily Johnson, Kate Glen, and Kimberly Pearman.
- Thank you to Ben Myatt, PharmD and DCA Pharmacy for compounding the material for Trial 1.
- Thank you Kris Fishman, Eric Huynh, and the team at Wells Pharmacy for compounding Niagen®Plus and helping identify a pathway to supply the material for Trial 2.
- Thank you, Chris Meletis, ND for reviewing the blood biomarker and vitals data.
- Thank you Albert Rembert for supervising clinical site visits and providing quality assurance and control guidelines.
- Thank you, Anthony Low, Esq., for taking care of all of the legal agreements to make the studies happen. You are such a valuable team member.
- Thank you Niagen Bioscience TruCrew and ELT for supporting our external research program and Raising the BAR.

## Abbreviations and Acronyms

ADP: adenosine diphosphate
ALT: alanine aminotransferase
AST: aspartate aminotransferase
Bili total: bilirubin total
BMI: body mass index
BUN: blood urea nitrogen
CI: confidence interval
CMP: comprehensive metabolic panel
DBP: diastolic blood pressure
DPN: diphosphopyridine nucleotide
ESR: erythrocyte sedimentation rate
FAS: fatigue assessment scale
FDA: Food & Drug Administration
GCP: good clinical practice
HHS: Health and Human Services
HR: heart rate
hsCRP: high sensitivity C-reactive protein
ID: identification
IM: intramuscular
IRB: institutional review board
ITT: intent to treat
IV: intravenous
LD50: lethal dose 50% population
MCH: mean corpuscular hemoglobin
MCHC: mean corpuscular hemoglobin concentration
MCV: mean corpuscular volume
NA: niacin
NAD+: nicotinamide adenine dinucleotide (NAD+)
NADH: reduced nicotinamide adenine dinucleotide NAM nicotinamide
NIH: National Institutes of Health
NMN: nicotinamide mononucleotide
NOAEL: no adverse events level
NR: nicotinamide riboside chloride
NRI: Nutraceuticals Research Institute
OHRP: Office for Human Research Protections
PARP: poly(ADP-ribose) polymerase
PI: primary investigator
RBC: red blood cell
RR: respiratory rate
SBP: systolic blood pressure
SC: subcutaneous
SpO2: peripheral capillary oxygen saturation
Temp: Temperature
WBC: white blood cell

